# Early Time-Restricted Eating Reduces Fat Mass In Morning Chronotype Adults With Overweight/Obesity

**DOI:** 10.64898/2026.11.17.26368370

**Authors:** Nicholas E. Phillips, Friedrich C. Jassil, Loic Metz, Andrew D. Biancolin, Julie Mareschal, Flore Sinturel, Céline Joris, Juan Sanchez-Avila, Alexandra Hemmer, Oksana Fiammingo, Stéphanie Blanc, Dylan Gendre, Marcel Salathé, Jardena J. Puder, Charna Dibner, Tinh-Hai Collet

## Abstract

Time-restricted eating (TRE) is a promising strategy to improve weight management and metabolic health, but its efficacy may vary depending on the coherence between the eating window and individual chronotype. In a randomized 12-week intervention of morning chronotype adults with overweight or obesity, early TRE (06:00h–14:00h) led to greater reductions in body fat mass compared to late TRE (12:00h–20:00h) and active control (three meals per day). Continuous glucose monitoring revealed enhanced circadian glucose rhythmicity and increased glycemic variability in early TRE, whereas postprandial glucose excursions were attenuated in late TRE. Early TRE induced the most pronounced systemic multi-omic changes, characterized by distinct transcriptional and metabolic programs, including reduced triglyceride and increased sphingomyelin levels. We found that in morning chronotype, early TRE and late TRE elicit distinct adaptations in body composition, glucose and lipid metabolism, and altered transcriptional and metabolic signatures.

*ClinicalTrials.gov registration no. NCT04618133*.

**HIGHLIGHTS:**

- A personalized window of meal timing is beneficial for metabolic improvement.
- Early time-restricted eating (e-TRE) enhances fat mass loss in morning chronotype.
- In e-TRE, meal timing impacts both circadian and postprandial glucose excursions.
- e-TRE leads to stronger reshaping of systemic metabolic and transcriptional programs.

**eTOC:** The benefits of early vs. late time-restricted eating (TRE) remain inconsistent across studies, though its efficacy may depend on meal timing relative to chronotype. We integrated cardio-metabolic assessment, high-resolution wearable data and multi-omics to demonstrate the differential effects of TRE timing on body composition, glucose dynamics, and blood multi-omic profiles.

## INTRODUCTION

Obesity increases the risk of non-communicable diseases and shortens life expectancy.^1^ Weight loss prevents and reduces these risks,^2,3^ but access to the most effective treatments (metabolic bariatric surgery and incretin-based medication) can be difficult due to limited availability, high costs or adverse effects.^4,5^ In addition, their efficacy also requires dietary and physical activity-related behavioral changes. Lifestyle modification thus remains the pillar of treatment for obesity management.^6^

In recent years, intermittent fasting has gained recognition as an effective dietary strategy for weight loss and metabolic health.^7,8^ Time-restricted eating (TRE) is a widely adopted form of intermittent fasting that alternates between a daily, restricted eating window and a fasting period aligned with the 24h cycle. However, TRE trials including adults with overweight or obesity have produced mixed results.^9–13^ This may be due to the lack of clear terminology^14^ and the heterogeneity across TRE implementations, particularly in the duration and the timing of eating window.^15,16^ Furthermore, the difficulty in adhering to the TRE protocol due to socio-familial activities and environmental cues,^17^ and the potential mismatch between the timing of TRE and individual chronotype, may also explain the inconsistent findings across studies.^18,19^

The circadian clock is an internal timing system that aligns physiology and metabolism with the daily changes of the environment at the individual and cellular levels.^20,21^ Chronotype refers to an individual’s intrinsic circadian preference for activity and optimal functioning at certain times of the day.^22^ Whether an individual is a morning (‘lark’), evening (‘owl’), or intermediate (neutral) chronotype is determined by a combination of biological (e.g., genetic, age, sex), environmental (e.g., light exposure, day length), and social factors (e.g., work/school timing that in turn impacts the timing of exercise and food).^23–25^ Individual differences in chronotype are mirrored at the cellular level as the circadian period length measured in skin fibroblasts of early chronotypes tends to be shorter than that of late chronotypes.^26^

In individuals with a morning chronotype, calorie consumption occurs earlier in the day, while individuals with an evening chronotype tend to eat later in the day.^25^ Because humans are diurnal, our metabolism is optimized for food intake earlier in the day,^27–29^ which may partly explain the increased cardiometabolic risk in evening chronotype.^30^ Accumulating evidence has shown greater benefits of TRE when the eating window is implemented early in the day.^31–33^ However, RCTs comparing the health benefits of early TRE (e-TRE) versus late TRE (l-TRE) in adults with overweight and obesity have yielded inconsistent findings,^10,34–40^ motivating further research to delineate their comparative metabolic effects. Importantly, it remains unknown whether personalizing the timing of TRE to the individual’s chronotype could maximize its health benefits.^41^

Wearable technologies enable continuous, high-temporal-resolution monitoring of behavioral and physiological real-world data, providing opportunities to refine lifestyle interventions.^42^ Smartphone food diary apps facilitate convenient recording of meal timing and content,^43–45^ providing insights into eating behavior in free-living conditions. Continuous glucose monitoring (CGM) records the 24- hour glucose profiles required to distinguish underlying circadian glucose rhythmicity from postprandial glucose excursions. We recently developed a computational model that integrates timestamped ingestion events with CGM data to represent glucose levels as the sum of postprandial excursions and an underlying 24h rhythm at the individual level.^46^ Whether these high-resolution wearable-derived readouts can identify novel biomarkers or provide mechanistic insights into metabolic adaptations to TRE remains to be established. In parallel, advances in metabolomics, lipidomics, and transcriptomics have improved our understanding of how diet affects the risk of non-communicable diseases.^47^ Circulating lipids and metabolites exhibit circadian rhythms that are altered in metabolic diseases.^48–50^ However, omics approaches have been applied infrequently and largely in isolation in TRE trials.^49,51–54^ Integrating multiple blood omics layers could reveal coordinated metabolic reprogramming according to TRE timing.

Our improved understanding of the pathophysiology of obesity has shifted the treatment paradigm from focusing on weight loss *per se* to targeting reduction of specific body compartments, i.e. body fat and visceral fat mass, while minimizing the loss of lean and bone mass.^55,56^ Our study therefore evaluated whether personalizing TRE timing to the individual’s chronotype could reduce body fat mass and optimize metabolic health in adults with overweight or obesity. We assessed clinical outcomes, wearable biomarkers, and multi-omic data by testing aligned vs. misaligned mealtimes with the morning chronotype, i.e. by comparing the effects of e-TRE (eating 06:00–14:00) vs. l-TRE (eating 12:00–20:00) vs. active control (three meals per day) for 12 weeks. e-TRE resulted in greater reductions in body weight and fat mass than l-TRE or active control. In CGM, e-TRE enhanced circadian glucose rhythmicity and increased glycaemic variability, whereas l-TRE attenuated postprandial glucose excursions. Multi-omics integration revealed that e-TRE coordinately reshaped the serum lipidome (lower triglycerides, higher sphingomyelins) and the monocyte transcriptome (reduced oxidative phosphorylation and fatty acid metabolism programs), consistent with rewired energy metabolism. Together, these findings reveal that TRE timing differentially regulates body composition, circadian glucose rhythmicity, and metabolic signaling networks among morning chronotype adults with overweight or obesity, providing insight into how alignment between meal timing and circadian clocks can be leveraged to improve metabolic health.

## RESULTS

### Study design

In a three-arm, open-label RCT, we recruited adults aged 25–50 years with a morning chronotype, body mass index (BMI) of 25–34 kg/m^2^ and an eating window lasting ≥ 12 hours. After checking eligibility criteria (**Methods**), all 92 enrolled participants underwent baseline assessment at visit 1, including questionnaires and fasting blood sampling (8–9 a.m.), followed by two weeks of wearable device monitoring. Key inclusion criteria were verified again after two weeks at visit 2: eating duration ≥ 12h, confident use of the smartphone app, and stable body fat mass within ± 1 kg (**Figure 1A**).

**Figure 1.**
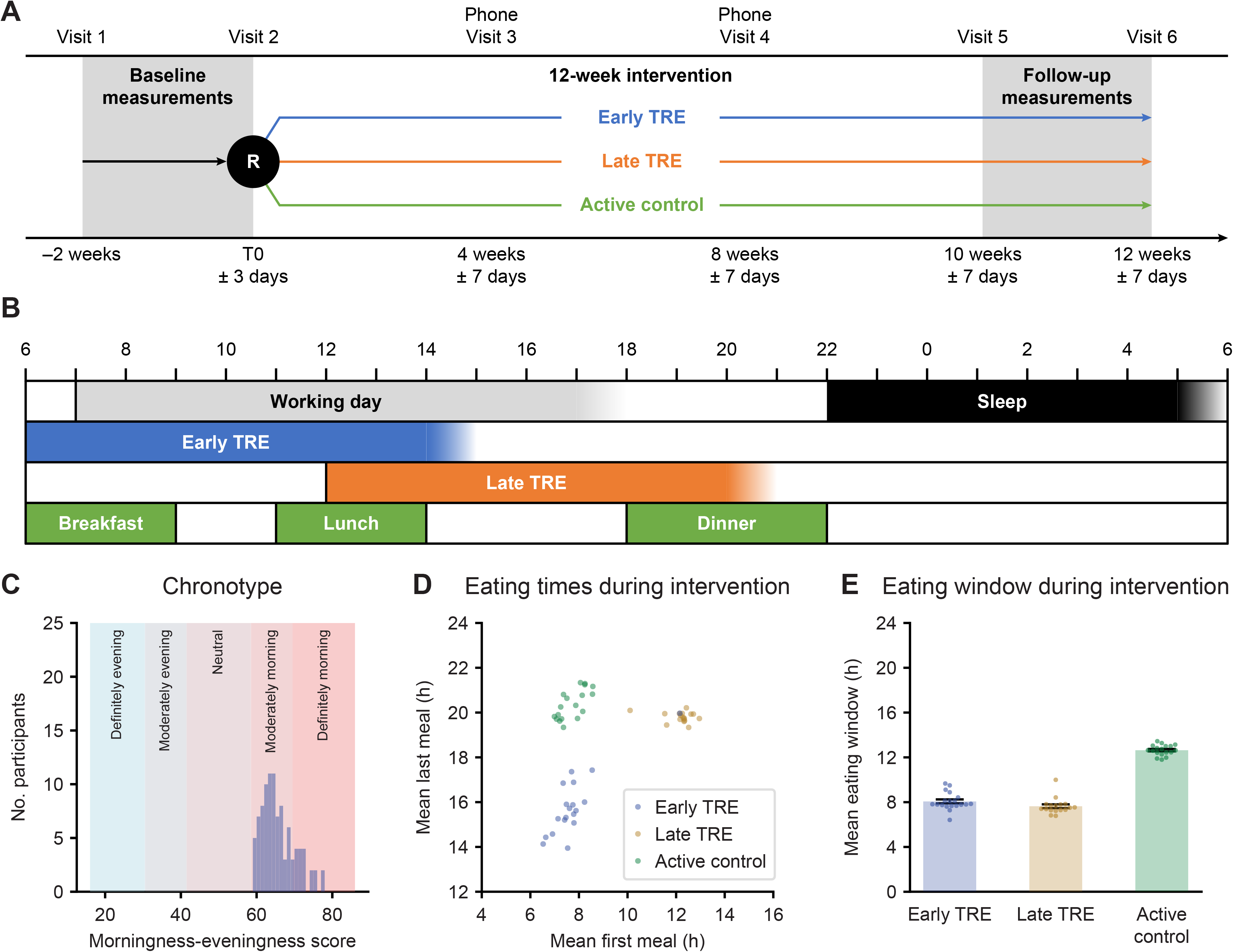
Overview of the study design and intervention adherence. (A) Baseline measurements including clinical assessments, questionnaires, and fasting blood samples at 8–9 a.m were performed at visits 1 and 2. After verification of key inclusion criteria at visit 2, participants were randomized (R) to one of the three intervention groups (see next panel). Data collection was repeated over the last two weeks of the 12-week long randomized intervention, i.e. at visits 5-6. (B) Participants were instructed to eat during an 8-hour window from 06:00h to 14:00h in the early TRE group (blue) and from 12:00h to 20:00h in the late TRE group (orange), with a 1-hour allowance according to their daily routine. In the active control intervention (green), participants were instructed to eat 3 meals per day (breakfast from 06:00h to 09:00h, lunch from 11:00h to 14:00h, dinner from 18:00h to 22:00h). (C) The distribution of chronotypes using the Horne-Östberg Morningness-Eveningness Questionnaire at baseline.^22^ (D) The mean timing of the first and last calorie intake for study participants during the intervention (visit 2 to 6), and (E) the mean duration of the eating window (time elapsed between first and last calorie intake) during the intervention period, as measured with the smartphone food diary app MyFoodRepo.^43–45^ <u>Abbreviations</u>: h, hours; R, randomization; TRE, time-restricted eating.

The 57 eligible participants were then randomized to e-TRE (eating 06:00h–14:00h) vs. l-TRE (eating 12:00h–20:00h) vs. active control (three meals per day) for 12 weeks (**Figure 1B**). Two participants withdrew from the study during follow-up and one was randomized by error (details in the **Methods**, **Figure S1**). A total of 54 participants thus completed the 12-week randomized intervention (visit 2 to 6) and are reported thereafter.

### Study population

Baseline characteristics of participants are presented in **Table 1** (and related **Table S1**). At baseline, participants had a mean age of 39.6 ± SD 6.7 years and 88% were women. The mean BMI was 29.7 ± 2.5 kg/m^2^, with 45 individuals (49%) in the overweight range and 47 (51%) in the obesity range. All had a morning chronotype by study design, with a median Horne-Östberg score of 64 (IQR 62–68, range 59–77), with no difference between randomization groups and 81.5% in the moderate morning group (**Table 1** and related **Table S1**, **Figure 1C**).

**Table 1.** Baseline characteristics of all enrolled participants (left column) and by randomized intervention group (right columns). Note that the final numbers of analyzed participants per intervention varied slightly due to study withdrawals or device-related technical issues (see text and Methods). Data are represented as mean ± SD, median (interquartile range) or number (%) as appropriate. ^1^ Chronotype was assessed with the MEQ by Horne and Östberg.^22^ ^2^ Body composition was assessed by DXA in 20 out of 21 participants of the early TRE group at baseline, due to limited availability of DXA measurement within the required timeframe. Some of the participants who did not meet all eligibility criteria before randomization (see **Figure S1**) declined the DXA measurement. <u>Abbreviations</u>: BP, blood pressure; CV, coefficient of variation; DXA, dual energy X-ray absorptiometry; MAGE, mean amplitude of glycemic excursions; MEQ, Morningness-Eveningness Questionnaire; TRE, time-restricted eating.

| Baseline characteristics | All enrolled | Randomized |  |  |
|---|---|---|---|---|
|  |  | Early TRE | Late TRE | Active control |
| **Demographics** | **(n = 92)** | **(n = 21)** | **(n = 17)** | **(n = 19)** |
| Age, years | 39.6 ± 6.7 | 38.9 ± 6.0 | 40.6 ± 6.8 | 39.8 ± 6.9 |
| Women | 74 (80%) | 17 (81%) | 16 (94%) | 17 (89%) |
| Chronotype (MEQ)<sup>1</sup> |  |  |  |  |
| Moderately morning | 75 (81.5%) | 18 (85.7%) | 12 (70.6%) | 16 (84.2%) |
| Definitely morning | 17 (18.5%) | 3 (14.3%) | 5 (29.4%) | 3 (15.8%) |
| **Clinical measurements** | **(n = 92)** | **(n = 21)** | **(n = 17)** | **(n = 19)** |
| Body weight, kg | 83.6 ± 10.7 | 80.1 ± 10.1 | 82.6 ± 9.4 | 82.9 ± 9.3 |
| Body mass index, kg/m<sup>2</sup> | 29.7 ± 2.5 | 29.8 ± 2.5 | 29.5 ± 2.4 | 29.9 ± 2.3 |
| Waist circumference, cm | 94.1 ± 7.8 | 94.1 ± 7.4 | 91.7 ± 7.6 | 94.2 ± 7.6 |
| Hip circumference, cm | 109.3 ± 8.0 | 107.2 ± 7.3 | 111.5 ± 7.4 | 111.0 ± 6.8 |
| Waist to hip ratio | 0.86 ± 0.07 | 0.88 ± 0.06 | 0.82 ± 0.05 | 0.85 ± 0.07 |
| Systolic BP, mmHg | 124.0 ± 13.9 | 124.0 ± 10.2 | 120.2 ± 11.3 | 126.5 ± 16.4 |
| Diastolic BP, mmHg | 77.0 ± 10.7 | 77.4 ± 10.8 | 74.8 ± 8.4 | 78.8 ± 12.1 |
| **Body composition by DXA<sup>2</sup>** | **(n = 81)** | **(n = 20)** | **(n = 17)** | **(n = 19)** |
| Fat mass, kg | 33.35 ± 5.54 | 32.02 ± 6.27 | 34.36 ± 4.68 | 33.04 ± 4.87 |
| Fat mass % | 41.3 (38.3 to 44.2) | 41.9 (37.0 to 44.3) | 40.4 (39.1 to 44.3) | 41.3 (39.0 to 42.7) |
| Visceral fat mass, g | 454.4 ± 161.0 | 540.5 ± 145.3 | 387.3 ± 149.4 | 386.9 ± 144.3 |
| Lean mass, kg | 46.93 (43.06 to 51.42) | 44.13 (42.05 to 52.01) | 46.12 (42.92 to 50.26) | 46.47 (43.45 to 50.19) |
| Bone mass, kg | 2.22 (1.99 to 2.58) | 2.00 (1.83 to 2.24) | 2.22 (1.98 to 2.46) | 2.36 (2.11 to 2.54) |
| **CGM metrics** | **(n = 86)** | **(n = 19)** | **(n = 15)** | **(n = 18)** |
| Mean glucose, mmol/l | 4.87 ± 0.55 | 4.80 ± 0.44 | 4.83 ± 0.45 | 4.78 ± 0.62 |
| CV, % | 14.83 ± 2.92 | 14.28 ± 2.34 | 15.53 ± 3.11 | 14.79 ± 2.90 |
| MAGE, mmol/l | 1.34 (1.13 to 1.55) | 1.28 (1.05 to 1.53) | 1.38 (1.15 to 1.59) | 1.28 (1.19 to 1.53) |

### Study participants followed the allocated meal timing

Participants recorded consumed food and beverages during the two weeks before the randomized intervention (visit 1 to 2) and the last two weeks of the 12-week intervention phase (visit 5 to 6, **Figure 1A**) using the smartphone food diary app MyFoodRepo.^43–45^ Timestamped calorie and macronutrient intake records thus allowed assessment of adherence to the allocated intervention (**Methods**).

The timing of the first and last meals revealed three defined groups according to the intervention (**Figure 1D**), with one participant switching intervention from e-TRE to l-TRE (details in the **Methods**). The mean time of the last meal in the e-TRE group (15:50h ± 01:25h) was later than the target of 14:00h (**Figure 1D**), indicating that participants had difficulty adhering to such an early eating cutoff. The mean duration of the eating window (i.e. the time elapsed between first and last calorie intake) was nonetheless 08:05h ± 00:50h in e-TRE and 07:35h ± 00:40h in l-TRE (**Figure 1E**), showing that both TRE groups attained the target restricted eating window of 8 hours, even if the e-TRE timing was slightly later than intended.

The eating window in the active control group (12:35h ± 00:30h) was similar to the mean baseline value of 12:50h ± 00:55h across all participants (p = 0.65, **Figure S2**), consistent with the maintenance of a similar eating window during the intervention. Participants were permitted one day per week without restrictions on meal timing, allowing greater flexibility for social and family activities.

### Early TRE led to greater body fat reduction compared to the late TRE or active control interventions

After 12 weeks, participants in the e-TRE group showed the greatest reduction in both absolute (kg, **Figure 2A**) and relative body weight (%, **Figure 2E**), with mean changes of –5.5% ± 4.2% with e-TRE, –1.6% ± 2.8% with l-TRE, and –0.6% ± 2.0% with active control (**Table S2**). The primary outcome of the study was the difference in body fat mass assessed with dual-energy X-ray absorptiometry (DXA). Within groups, body fat mass decreased by a median of -2.90 kg (IQR – 1.74 to –3.61; p < 0.001) with e-TRE and -1.37 kg (IQR, −0.24 to 2.16; p = 0.045) with l-TRE, but remained unchanged with active control (−0.02 kg, IQR, −1.21 to 1.63; p = 0.93; **Figure 2B**). In group comparisons, participants in the e-TRE group lost significantly greater body fat mass compared to both active control (–3.04 kg, 95%CI –4.78 to –1.29 kg, p = 0.001) and l-TRE groups (–1.93 kg, 95%CI –3.72 to –0.14 kg, p = 0.035) (**Figure 2I, 2K**), which was also observed for fat mass measured with bioelectrical impedance analysis (BIA, **Table S2**). In contrast, the change in body fat mass was not significantly different between l-TRE and active control groups (**Figure 2J**, **Table S2**).

**Figure 2.**
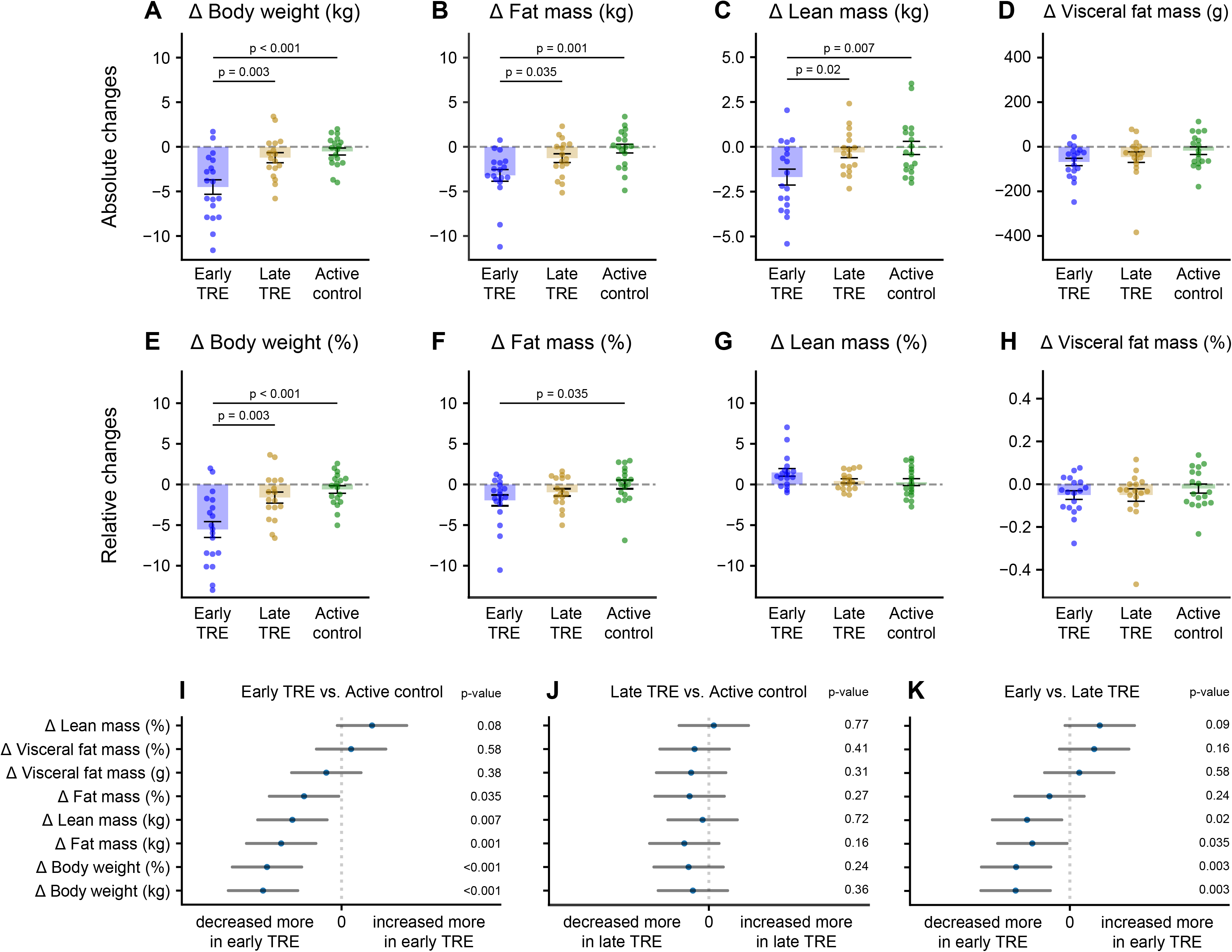
Early TRE led to greater loss of body weight, fat mass, and lean mass than late TRE or active control interventions. (A–D) Change in absolute terms (kg) from pre- to post-intervention of (A) body weight measured with a calibrated scale, (B) fat mass, the primary outcome, (C) lean mass, and (D) visceral fat mass, measured by dual-energy X-ray absorptiometry. Each dot represents an individual participant, shown separately for each group. (E) Body weight percentage change after 12 weeks relative to baseline (%) and (F-H) represent body composition outcomes as a percentage of total body weight (%). (I–K) Pairwise group comparisons for key body composition outcomes, calculated using ANCOVA. Dots indicate regression coefficients representing group differences, and lines show 95% confidence intervals. P-values are shown on the right of each panel. <u>Abbreviations</u>: ANCOVA, analysis of covariance; TRE, time-restricted eating.

The e-TRE group also lost a median of –1.8 kg (IQR –3.2 to –0.2 kg) of lean mass (**Figure 2C**), which was greater than both the active control (p = 0.007) and l-TRE groups (p = 0.02, **Figure 2I, 2K**), and supported by similar results for fat-free mass as measured with BIA (**Table S2**). However, there were no statistically significant changes between groups when lean mass was considered as a percentage of body weight (**Figure 2G**, **Table S2**). Visceral fat mass was significantly reduced only in e-TRE (median –48.0 g, IQR –106.8 to –20.8 g; p < 0.001; **Figure 2D**), but between-group differences were not significant (**Figures 2I–K**), either in absolute terms or when expressed as a percentage of total body weight (**Figure 2H**). In an additional *per protocol* analysis (i.e. accounting for one participant who switched from e-TRE to l-TRE group), we found similar results.

### Intervention groups differed significantly in macronutrient consumption patterns

The smartphone food diary app MyFoodRepo captures detailed changes over the 24h cycle in dietary timing and composition, i.e. calories, carbohydrates, protein, fat (**Figure 3**), fibre, and alcohol (**Figure S3**), which allowed detailed comparison of macronutrient consumption patterns between baseline (visit 1 to 2) and the intervention phase (visit 5 to 6). The analysis of calorie intake confirmed that participants followed their allocated intervention: the e-TRE group had a large and significant decrease during the evening period, the l-TRE group showed reduced intake during the morning period, and the three-meal-per-day active control group showed a significant decrease during the mid-afternoon snack period (**Figure 3A**). The mean number of calories consumed per day changed by –399.3 ± 286.0 kcal/d (p < 0.001) in the e-TRE group, –196.7 ± 239.1 kcal/d (p = 0.005) in the l-TRE group, and –132.7 ± SD 369.0 kcal/d (p = 0.16) in the active control group. The calorie reduction was nominally greater with e-TRE than with l-TRE or active control (both unadjusted p = 0.02), but these differences did not remain statistically significant after correction for multiple testing across all nutritional outcomes (**Figure 3E**, **Table S4**).

**Figure 3.**
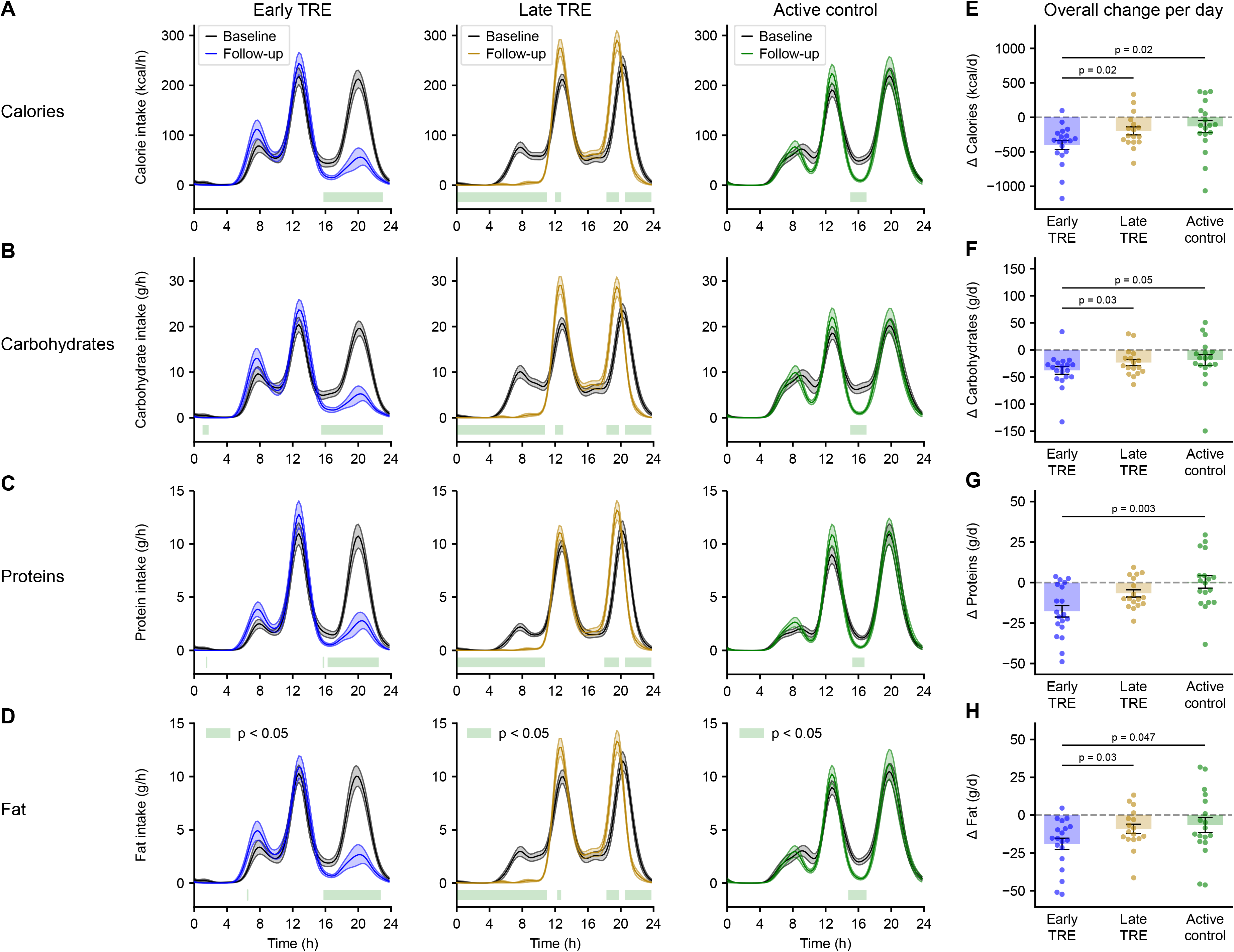
Food diary recordings show reduced evening-meal intake in the early TRE group and reduced breakfast intake in the late TRE group across calories, carbohydrates, proteins, and fat. (A-D) The mean consumption rate per hour for the early TRE (blue), late TRE (orange) and active control group (green), for (A) total calories, (B) carbohydrates, (C) proteins, and (D) fat at baseline (black line) and follow-up periods (colored line). The shaded area represents the mean ± SEM, and the green bar at the bottom represents significant differences from baseline, as assessed from p < 0.05 in a Wilcoxon non-parametric test, adjusted for multiple testing with the Benjamini-Hochberg procedure. (E-H) Individual change in mean daily intake for early TRE (blue), late TRE (orange) and active control (green) for total calories, carbohydrates, proteins, and fat, respectively. <u>Abbreviations</u>: d, day; h, hours; TRE, time-restricted eating.

The global temporal pattern of calorie intake (**Figure 3A**) was similarly observed for carbohydrates (**Figure 3B**), protein (**Figure 3C**), fat (**Figure 3D**) and fibre (**Figure S3A**) in each intervention. The evening meal was overall a richer source of protein, fat and fibre than breakfast (**Figure 3C-D**, **Figure S3A**), and the strongest daily macronutrient changes were observed in the e-TRE group (all p < 0.001, **Figure 3F-H**, **Figure S3C**). The protein decrease in e-TRE (from 76.4 ± 17.5 to 58.4 ± 16.9 g/day) remained within physiologically safe limits, but may need to be accounted for in clinical decision-making for specific patient populations (see **Discussion**). There were no significant changes in alcohol consumption for any of the intervention groups (**Figure S3B**, **S3D**).

To explore whether the observed effects of TRE were attributable to changes in energy intake vs. meal timing, we examined the correlations between weight change and concurrent changes in calorie intake and eating window duration. Across all participants, calorie reduction correlated modestly with weight loss (Pearson ρ = 0.28, 95%CI 0.02 to 0.51, p = 0.04, **Figure S4A**), as did the shortening of eating window (ρ = 0.32, 95%CI 0.06 to 0.54, p = 0.02, **Figure S4B**). There was no association between changes in eating window and calorie intake (p = 0.13, **Figure S4C**). In a sensitivity analysis adjusting for the change in calorie intake, the body fat mass loss (i.e. the primary outcome) remained significantly greater in e-TRE than both l-TRE (p = 0.026) and active control (p = 0.002). However, the relative contributions of calorie reduction and eating window shortening to overall loss of body weight and fat mass are difficult to quantify in this pragmatic, real-world trial where calorie intake was not strictly controlled and measured.

### Early and late TRE associated with differential changes in glycemic variability

We used CGM for 2 weeks at baseline (visit 1 to 2) and during the intervention (visit 5 to 6) to assess the impact of TRE on glucose metabolism. No significant changes were observed for mean glucose in either group when averaged across the entire CGM recording period (**Figure 4A**). Considering the whole measurement period, e-TRE led to greater glycemic variability, with increased CV relative to both active control (2.6%, 95%CI 1.1 to 4.1%, p = 0.001) and l-TRE (2.9%, 95%CI 0.7 to 5.1%, p = 0.01, **Figure 4B**). Similarly, MAGE was higher after e-TRE compared with the l-TRE and active control groups (both p ≤ 0.003, **Figure 4C**), but the absolute differences were small in clinical terms (**Table S5**). These differences remained statistically significant after correction for multiple testing.

**Figure 4.**
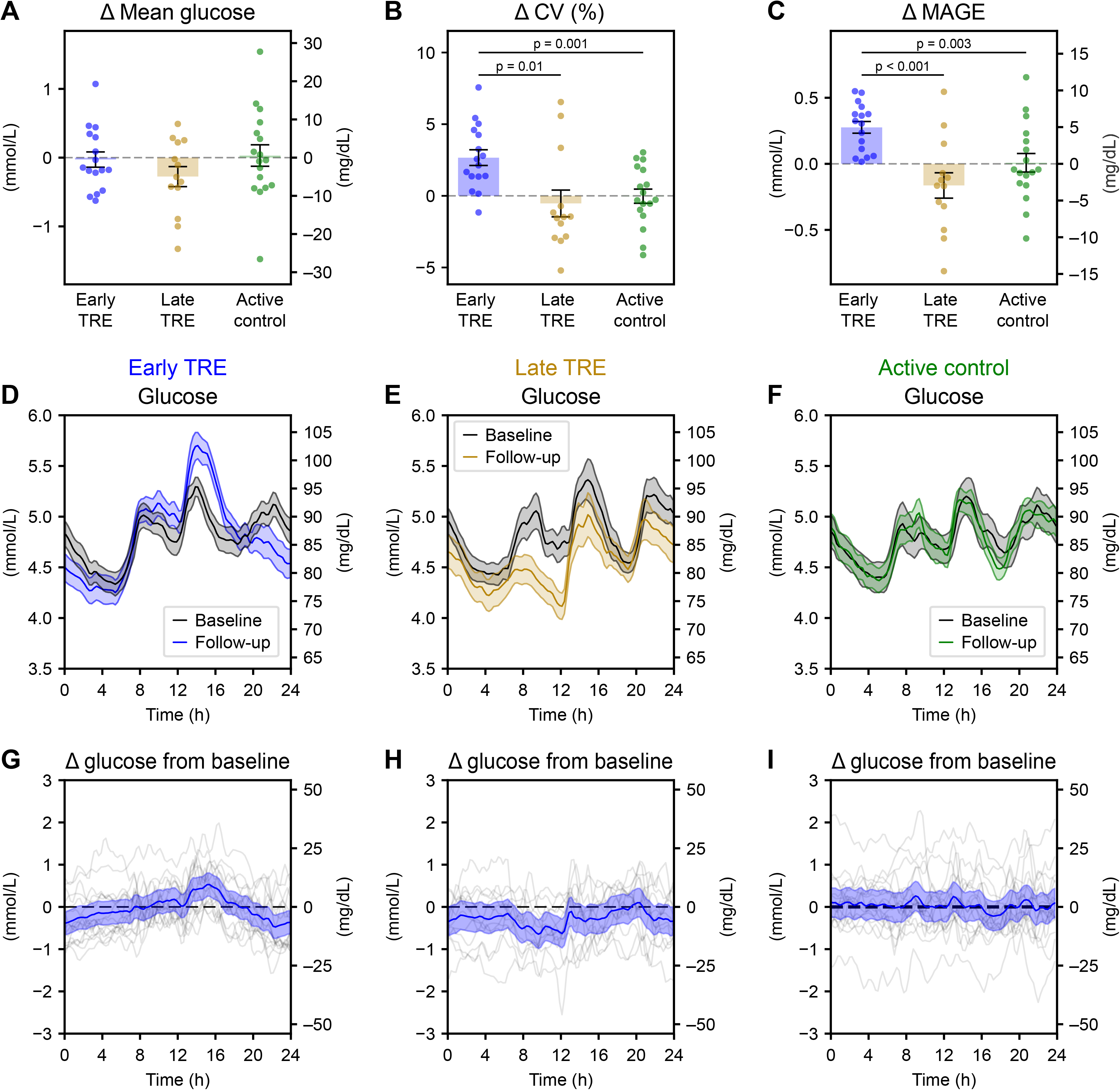
Changes in glycemic mean, variability and 24-hour glucose profile curves, as measured with CGM. (A-C) Change from pre- to post-intervention of mean glucose levels, CV and MAGE, respectively, using CGM data across the 2-week measurement period. (D-F) The 2-hour moving average estimate over the 24-hour clock of mean glucose levels for the control, early TRE, and late TRE interventions, respectively. The shaded area represents the mean ± SEM. (G-I) Change in mean glucose levels from pre- to post-intervention. Each participant is represented by a grey line, while the group mean (blue) and 95% confidence interval (shaded blue) are derived from a linear mixed-effects model. <u>Abbreviations</u>: CGM, continuous glucose monitoring; CV, coefficient of variation; MAGE, mean amplitude of glycemic excursions; TRE, time-restricted eating.

CGM further allowed estimation of how mean glucose levels varied over the 24-hour cycle in each group (**Figure 4D-F**). Based on linear mixed-effects modelling, the active control group showed no notable changes from baseline to follow-up (**Figure 4G**). With e-TRE, glucose levels were lower during the late evening (20:00h–00:00h), but higher by approximately 0.5 mmol/l in the afternoon (12:00h–16:00h, **Figure 4H**). Note that the higher afternoon glucose levels were not accompanied by strong differences in carbohydrate consumption at lunchtime compared to baseline behaviour (**Figure 3B**). The l-TRE group exhibited lower glucose levels of approximately 0.5 mmol/l in the morning (08:00h–12:00h, **Figure 4I**). Although this analysis captures overall 24-hour glucose patterns, it cannot disentangle the relative contributions of meal-driven postprandial responses vs. intrinsic circadian modulation of glucose levels, motivating the application of our recently published computational model.^46^

### TRE modulated underlying glucose circadian rhythms and postprandial excursion

We next applied a mathematical model of glucose dynamics, which represents glucose levels as the sum of two components^46^: (1) postprandial glucose excursions using the timestamps from the ingestion events, and (2) a 24-hour rhythm in baseline glucose levels (schematic in **Figure 5A**). In the e-TRE group only, the amplitude of baseline 24-hour glucose rhythms increased by a median of 0.28 (IQR 0.06 to 0.42) mmol/L compared to baseline, which was significantly greater than the active control group (p = 0.002, **Figure 5B, Table S5**). This amplitude increase, as illustrated by participant ID 57 in the e-TRE group (**Figure 5D-E**), reflects a pattern previously associated with a healthier metabolic profile.^57^ Conversely, the mean height of postprandial glucose excursions was lower in l-TRE compared to e-TRE by a mean of 0.20 mmol/L compared to baseline (95%CI 0.06 to 0.33, p = 0.008, **Figure 5C**, **Table S5**), as exemplified by participant ID 85 in the l-TRE group (**Figure 5F-G**).

**Figure 5.**
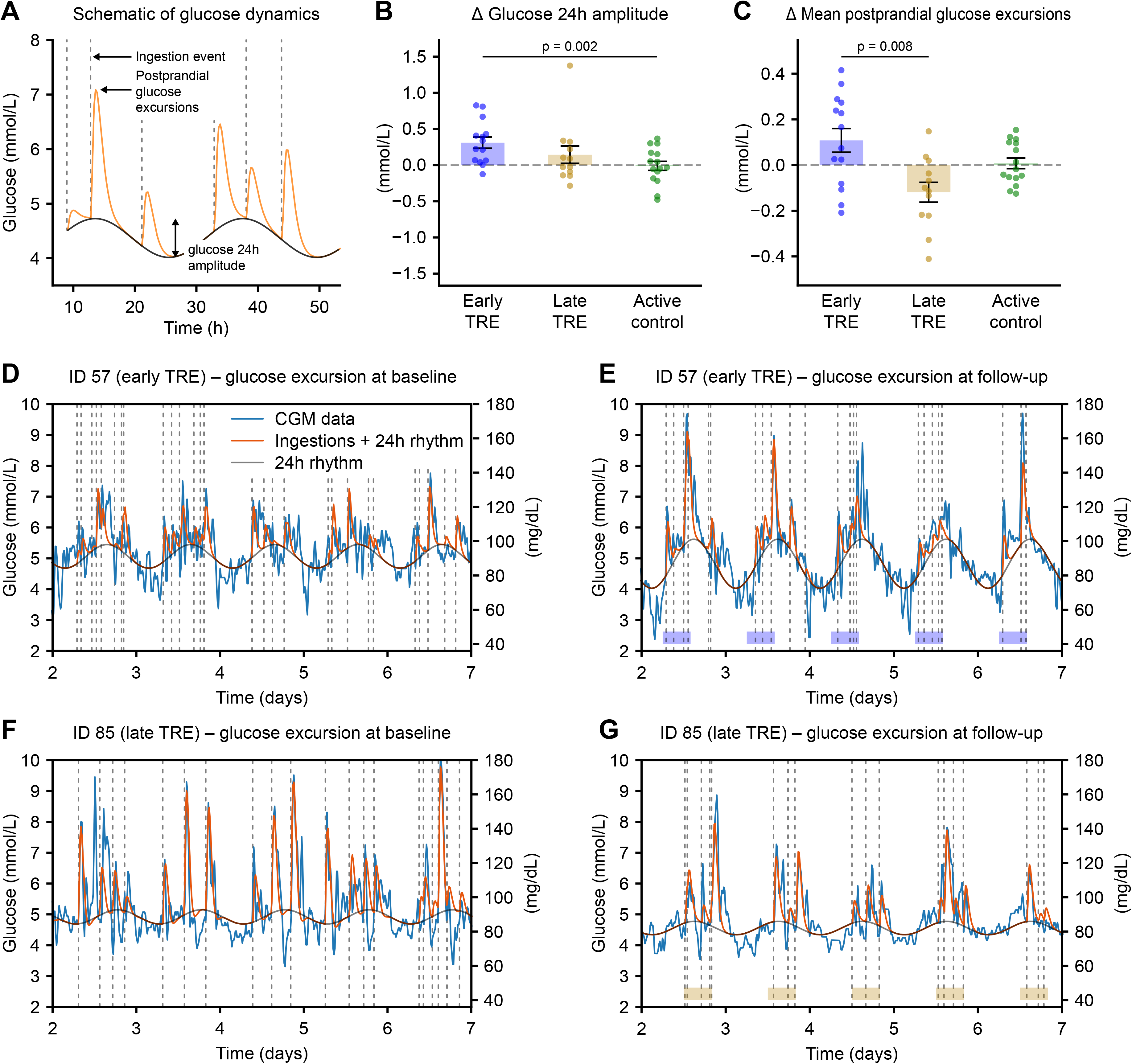
TRE alters circadian glucose rhythm and postprandial response. (A) Schematic of the mathematical framework, which uses meal events (dashed lines) to model glucose levels as a superposition of postprandial glucose excursions (orange) on top of an underlying 24-hour rhythm (black).^46^ (B) Changes from pre- to post-intervention of the glucose 24-hour amplitude. (C) Change from pre- to post-intervention of the mean postprandial glucose excursions. (D-G) Individual examples for the early TRE group (ID 57, D-E) and the late TRE group (ID 85, panels F-G) of CGM data and the corresponding model fit at baseline (D,F) and follow-up (E,G). The shaded area at the bottom (E,G) represents the allocated eating window (E: early TRE; G: late TRE). Blue, CGM raw data; orange, model prediction incorporating ingestion events and the underlying glucose 24-hour rhythm; black, the underlying glucose 24-hour rhythm; dashed lines, ingestion events. <u>Abbreviations</u>: CGM, continuous glucose monitoring; ID, identification code; TRE, time-restricted eating.

### Early TRE led to reduced fasting plasma glucose, insulin resistance and sleep duration

In addition, we assessed changes in metabolic health by clinical measurements, indirect calorimetry, and metabolic blood profile in the fasting state, as detailed in the **Supplementary Results**. Fasting plasma glucose decreased in e-TRE compared to active control (–0.27 mmol/l, 95%CI –0.52 to –0.03 mmol/l, p = 0.03), as well as insulin resistance (HOMA-IR: –0.57, 95%CI – 1.09 to –0.05, p = 0.03, **Table S3**). However, neither difference remained statistically significant after correction for multiple testing. Of note, the mean glucose levels measured with CGM over two weeks were not different across interventions (**Figure 4A**, **Table S5**).

While there was no change in self-reported physical activity, the actigraphy indicated a slight increase in moderate activity in the active control group (11.0, IQR 4.9 to 22.5 minutes), which was nominally greater than the e-TRE and l-TRE groups (both p ≤ 0.004, **Table S3**). The actigraphy-measured sleep duration was also nominally shorter in e-TRE vs. l-TRE (–0.42, IQR –0.78 to –0.06 hours, p = 0.02). However, none of these differences remained statistically significant after correction for multiple testing.

### TRE does not shift skin fibroblast clocks or internal circadian phase

Primary skin fibroblasts synchronized *in vitro* represent a good proxy for the internal clockwork.^58,59^ To determine if TRE could have an impact on molecular oscillators, we recorded circadian bioluminescence of *Bmal1*-luciferase reporter from primary skin fibroblasts established from skin biopsies collected in a subset of participants before and after intervention (**Figure S5**). Analyses of intrinsic circadian parameters suggested no substantial change in period, acrophase and amplitude from baseline in any intervention (all p > 0.05, **Figure S6C-E**).

To assess whether TRE shifted the internal phase, we estimated internal circadian time from the single-time point monocyte RNA-seq with two independent algorithms, TimeMachine^60^ and ZeitZeiger.^61^ Neither method detected internal phase shift after any nutritional intervention (all p > 0.05, **Figure S6F**), and this did not differ between groups (all p > 0.05). Per-subject shifts agreed between the two methods (Spearman ρ = 0.66, p < 0.001, **Figure S6G**). Overall, these data indicate that TRE is not substantially affecting circadian oscillations assessed either in primary skin fibroblasts synchronized *in vitro,* or in blood monocytes.

### Early TRE induced a coordinated systemic metabolic remodeling

To determine whether meal timing differentially alters systemic metabolism and the monocyte transcriptional state, we performed multi-omics profiling across circulating metabolites, lipids and monocyte transcripts pre- and post-intervention within the same group (**Figure S7**), between groups (**Figure S8**), as well as by an integrated approach (**Figure 6A**). A total of 13,051 transcripts were compared before and after nutritional intervention. e-TRE, but neither l-TRE nor active control, induced broad monocyte gene expression remodeling with 2057 differentially expressed genes following the intervention (FDR < 0.05, **Figure S7A-C**). Transcripts associated with metabolic regulation such as *RAI1*, the clock modulating nuclear receptor *RXRA,* and the sphingomyelin synthase 2 (*SGMS2*) were upregulated, alongside the core clock genes *PER2* and *CRY1.* On the other hand, 3 members of the S100A monocyte alarmins were downregulated (*S100A8, S100A9* and *S100A12*). Gene set enrichment analysis using the Molecular signature database (MSigDB) hallmark collection revealed significant changes only within the e-TRE group. The top 10 enriched sets included negative enrichment of oxidative phosphorylation and fatty acid metabolism, and positive enrichment of Wnt/β-catenin and Notch signaling (FDR < 0.05, **Figure 6B**). By contrast, comparison between groups revealed no differences at either the feature or the hallmark set level (**Figure 6B, Figure S8A-C**).

**Figure 6.**
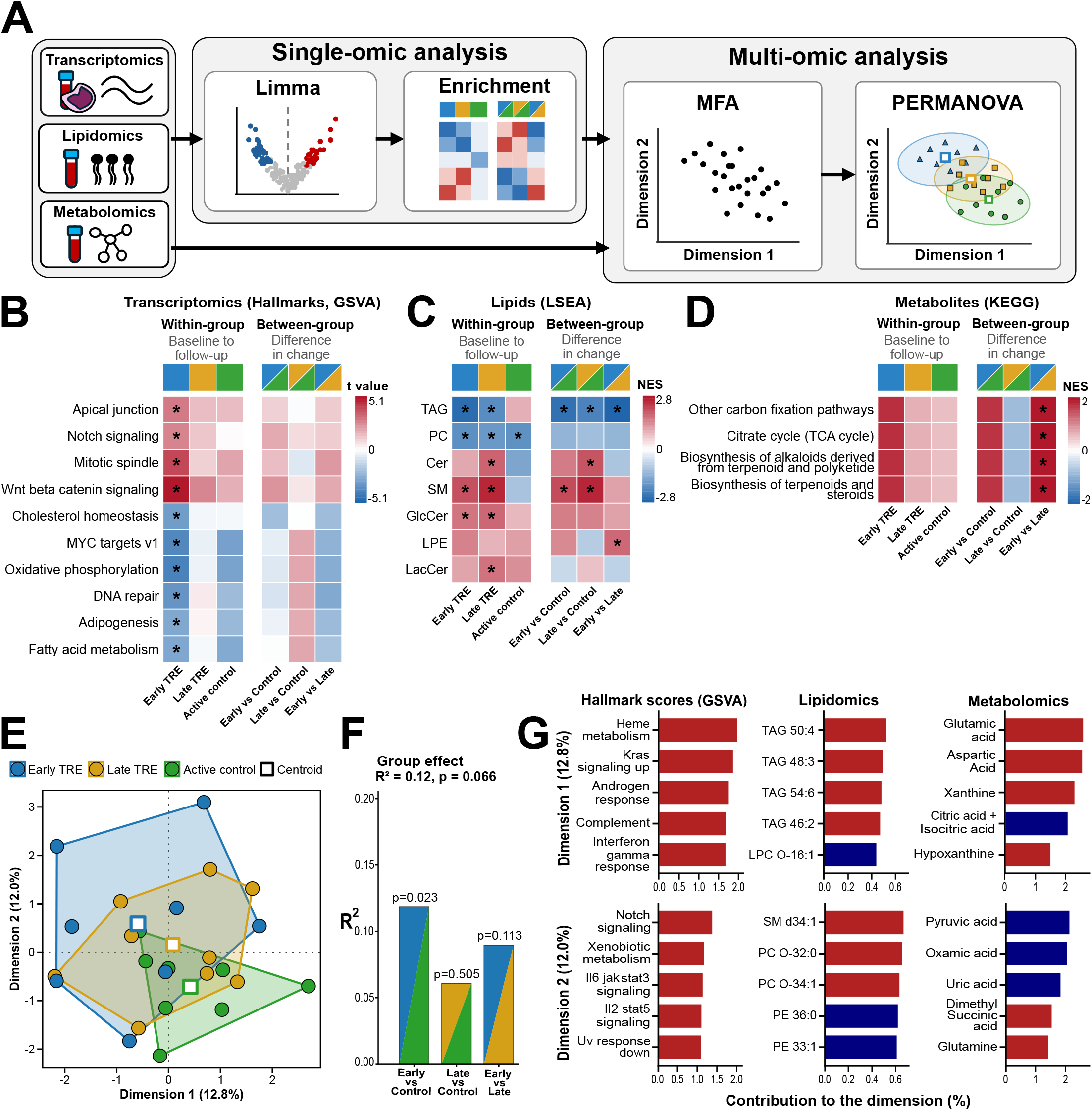
Pathways analysis and integrated multi-omic analysis identifies an early TRE molecular signature. (A) Analysis workflow. Transcriptomics (monocyte RNA-seq), lipidomics and metabolomics were each analyzed by limma and set enrichment for baseline to follow-up changes within and between regimens (single-omic analysis), and the changes from baseline were integrated by MFA and tested by PERMANOVA (multi-omic analysis). (B–D) Set-level changes in transcripts (B), lipids (C) and metabolites (D). For each layer, the left block shows within-group changes from baseline to follow-up in e-TRE (blue), l-TRE (orange) and active control (green). The right block shows between-group differences in change (e-TRE vs. control, l-TRE vs. control, e-TRE vs. l-TRE). Asterisks (*) mark FDR < 0.05. Sets with FDR < 0.05 in at least one contrast are shown, up to ten per layer. (E) MFA individual map of the Δbaseline to follow-up profiles of the three omics blocks (hallmark GSVA scores, lipids, metabolites). (F) PERMANOVA on the first ten MFA dimensions. (G) Variables contributing most to MFA dimensions 1 (top) and 2 (bottom), for each omic block. Bars show the contribution to the dimension (%). All omics analyses were performed in a subset of participants: total n = 25, e-TRE n = 8, l-TRE n = 8, and active control n = 9. <u>Abbreviations</u>: Cer, ceramides; GlcCer, glucosylceramide; GSVA, gene set variation analysis; KEGG, Kyoto encyclopedia of genes and genomes; LacCer, lactosylceramide; LPC O-, ether-linked lysophosphatidylcholine; LPE, lysophosphatidylethanolamine; LSEA, lipid set enrichment analysis; MFA, multiple factor analysis; PC, phosphatidylcholine; PC O-, ether-linked PC; PE, phosphatidylethanolamine; PERMANOVA, permutational multivariate analysis of variance; SM, sphingomyelin; TAG, triacylglycerol; TCA, tricarboxylic acid; TRE, time-restricted eating.

Of the 225 detected lipids species, l-TRE and active control exhibited no baseline-to-follow-up detectable changes in individual species, whereas e-TRE led to a change in the abundance of 42 lipid species (**Figure S7D-E**). Among those, several triglycerides (e.g. TAG 44:1, 44:2, 46:2) and phosphatidylcholines (e.g. PC 30:0, 34:5, 36:5) were less abundant, while sphingomyelins (e.g. SM d35:1, d36:1, 36:2) were more abundant after e-TRE. Indeed, lipid set enrichment analysis revealed an overall depletion of triglycerides (FDR < 0.001) and phosphatidylcholine (FDR = 0.002) species, alongside enrichment of sphingomyelins (FDR = 0.003) and glucosylceramides (FDR = 0.045) following e-TRE. A similar pattern was found with l-TRE, but not in the active control group. Yet, triglycerides as species (**Figure S8D-E**) and as class (**Figure 6C**) showed significantly greater reductions in e-TRE compared to l-TRE and active controls (FDR < 0.001).

Serum metabolomic analyses revealed neither significant changes in the 93 metabolites detected after any of the interventions (**Figure S7G-I**), nor differences among these changes across nutritional interventions (**Figure S8G-I**). At the same time, metabolite set enrichment analysis revealed that subjects following e-TRE had enriched TCA (tricarboxylic acid) cycle intermediates (e.g. succinate, citrate, α-ketoglutarate) from baseline to follow-up compared to l-TRE (FDR = 0.02, **Figure 6D**).

### Integrated multi-omic analysis suggests an early-TRE signature relative to active control

To determine whether these univariate differences translated into a systems-level signature of the nutritional intervention, we integrated the changes from baseline in Hallmark gene set enrichment scores, lipids and metabolites by multiple factor analysis (MFA) and tested group effects by permutational multivariate analysis of variance (PERMANOVA) (**Figure 6A**). The first ten dimensions explained 71.2% of the variance. Participants from the three groups overlapped on the first two dimensions, with e-TRE lying furthest from the active control along both axes (**Figure 6E**). PERMANOVA on the first ten dimensions showed trend toward a group effect (omnibus R^2^ = 0.12, p = 0.07), driven by the difference between e-TRE and active control (R^2^ = 0.12, p = 0.02, **Figure 6F**), whereas the l-TRE vs. active control (p = 0.51) and e-TRE vs. l-TRE (p = 0.11) contrasts were not significant. There was no difference in homogeneity of multivariate dispersion between regimens based on the PERMDISP (p = 0.33). Dimension 1 was driven by hallmark sets such as complement and interferon-γ response, triglyceride species and amino acid/purine metabolites (glutamic and aspartic acid, xanthine, hypoxanthine), whereas dimension 2 was dominated by IL2-STAT5 and IL6–JAK–STAT3 signaling, sphingomyelin and ether-linked phosphatidylcholine species, and pyruvate-related metabolites (pyruvic and oxamic acid) (**Figure 6G**). These multi-omic analyses are consistent with the single-block results (see previous section): e-TRE showed the most distinct systems-level profile relative to active control, with lipid (triglyceride and sphingomyelin species) and immune/transcriptional features among the main contributors, although the overall group effect did not reach significance.

## DISCUSSION

This study investigates the differential effects of TRE matched or not with individual chronotype, examined in adults with overweight/obesity and a homogenous morning chronotype. In agreement with our hypotheses, e-TRE leads to greater reductions of body weight and fat mass than the l-TRE and active control interventions. Early meal timing enhanced circadian glucose rhythmicity, but increased glycemic variability in CGM. Chronotype-aligned early, but not late, TRE was associated with coordinated changes in monocyte gene transcripts and in circulating lipids. Significant changes induced by e-TRE comprised a greater triglyceride reduction and increased circulating TCA cycle intermediates.

Body fat mass, the primary study outcome, decreased more with e-TRE than l-TRE or active control interventions after 12 weeks, in keeping with a recent RCT showing greater loss of fat mass when the timing of TRE is personalized to chronotype.^33,37,40^ It is nonetheless important to note that, in contrast to our trial, the previous study also prescribed caloric restriction to the TRE and control groups, explaining a significant loss of fat mass in both groups and making it difficult to isolate the effects of TRE without deliberate calorie restriction. Other RCTs comparing e-TRE and l-TRE showed no difference in fat mass, which could be due to a shorter duration, a combination with caloric restriction, a mismatch between mealtimes and chronotype, or TRE implementation.^34,36,38,53^

Data from the MyFoodRepo smartphone app confirmed that the TRE interventions effectively reduced the eating window to 8 hours. Although participants in e-TRE were permitted to extend the 14:00h limit to 15:00h, the observed mean last meal time of 15:50h ± 01.25h suggests that adhering to an early eating schedule may be challenging in free-living conditions. Collective data from 10 RCTs has also shown lower adherence and retention rates in e-TRE than l-TRE,^62^ where this difficulty potentially reflects a combination of physiological (e.g., hunger), social, and habitual factors, especially over multiple months.^18,63^ Although e-TRE markedly reduced late-day eating, participants did not substantially compensate for the lost calories at breakfast, resulting in a spontaneous reduction in daily calorie intake that was numerically greater than in l-TRE and active control, although these between-group differences did not remain significant after correction for multiple testing. Indeed, a decreased calorie intake in the absence of intentional efforts is presumably one of the mechanisms of weight loss following TRE interventions.^64^ Adjusting for changes in calorie intake did not alter the observed between-group differences in fat mass loss and hence supports an effect independent of calorie restriction, although this finding should be interpreted with consideration of potential measurement error in dietary reporting. The relative contributions of reduced calorie intake, eating-window timing and changes in physical activity to the observed weight loss remain challenging to precisely quantify in this real-world setting.

CGM revealed a distinct 24-hour glucose profile with e-TRE, characterized by lower nocturnal glucose, higher early afternoon glucose, and greater glycemic variability, without changes in mean glucose values. In computational modelling this variability arose from both an increased amplitude of 24-hour glucose rhythm and larger postprandial glucose excursions, despite lower carbohydrate intake. Interestingly, our previous work using the same modeling framework associated higher circadian amplitude with better, potentially more flexible, metabolic health, as it was greater in non-gestational diabetes than in gestational diabetes postpartum women.^57^ Although it contributes to glycemic variability, the increased amplitude observed in e-TRE may reflect a favourable metabolic adaptation in otherwise metabolically healthy subjects. By contrast, postprandial glucose excursions were reduced with l-TRE compared to e-TRE. Thus, while e-TRE was associated with stronger circadian glucose rhythms, l-TRE led to attenuation of postprandial glucose excursions.

The reduction in lean mass within and between groups followed the same trends as the loss of fat mass. As fat mass loss was 1.7-fold greater than lean mass loss in e-TRE, the change in the proportion of lean mass relative to total body weight did not significantly differ from other groups. Concurrent lean mass reduction is a common problem during any weight loss intervention.^65–67^ The e-TRE group also reduced protein, fat, and fibre intake compared to active control, which may not be advisable in all cases. Future strategies to mitigate the loss of lean mass during TRE could include resistance training, dietary protein optimization, or, where appropriate, pharmacological support for muscle maintenance.^65^ This would however increase the complexity of TRE implementation, possibly lowering the adherence rate.^18,62^

The multi-omic analyses extended the clinical and wearable findings by showing that e-TRE induced the most distinct lipidomic and monocyte transcriptomic signature compared to l-TRE, consistent with a previous study reporting lipid remodeling in e-TRE but not l-TRE led by different features.^54^ e-TRE decreased circulating triglycerides and increased sphingomyelins. The triglyceride decrease is consistent with the greater loss of fat mass^68,69^ and the total triglyceride reduction measured in clinical chemistry, whereas sphingomyelin responses to TRE timing or weight loss are heterogeneous.^54,69^ Therefore, the increase in sphingomyelins cannot be attributed to weight change alone. Monocytes contribute little to circulating sphingomyelins, but *SGMS2* was upregulated in e-TRE alongside higher plasma sphingomyelin, suggesting a parallel shift in sphingolipid metabolism. The fat mass loss and lower triglyceride pool also came with a larger decrease in monocyte fatty acid metabolism and oxidative phosphorylation programs, which may reflect lower substrate availability and reduced cellular energy metabolism as expected with weight loss.^51^ MFA showed that the multi-omic profile differed between e-TRE and active control, with the first two dimensions composed of triglycerides, immune-regulatory gene sets and TCA cycle-related metabolites, in line with the single-block results. Weight loss is often accompanied by lower low-grade inflammation and less pro-inflammatory monocyte programs.^70,71^

Whereas blood monocytes displayed a change in expression of core clock genes *PER2* and *CRY1* only after e-TRE, the estimation of internal circadian time derived from single-time point blood monocyte RNA-seq showed no phase shift. Furthermore, no significant changes were detected in circadian rhythms of primary skin fibroblasts, assessed following *in vitro* synchronization, across nutritional interventions. Of note, the individual rhythms evaluated by actigraphy stayed largely unchanged after TRE in both regimens, consistent with other studies.^72,73^ Thus, the observed expression changes of several clock genes such as *PER2*, *CRY1*, *RAI1* or *RXRA* could be attributed to their clock-unrelated metabolic roles rather than to their core-clock function. Previous work revealed that a 5-hour change in meal timing for seven days altered the acrophase of *PER2* expression in adipose tissue, with no corresponding changes in whole blood cells.^72^ Therefore, the effect of TRE on peripheral clocks might be tissue- and time-specific and may possibly only be detected in metabolically active tissues.

A major strength of this study is that it directly compared e-TRE and l-TRE in a homogenous morning chronotype population of adults with overweight or obesity.^37,38^ To study the TRE effect *per se* and in contrast to other studies,^34,37,38^ we purposely did not combine TRE with other dietary strategies such as calorie restriction or a Mediterranean diet. Second, we strictly followed the definition of morning chronotype at inclusion, ensuring that the TRE intervention matched or not the individual chronotype. Third, multiple wearable devices assessed the temporal changes in clinical parameters and CGM data to account for circadian variation across 24h. The MyFoodRepo smartphone app ensured detailed assessment of meal timing and macronutrient intake, enabling objective evaluation of adherence to the assigned nutritional intervention and postprandial glucose excursions when combined with the concomitant CGM data. Lastly, the embedded multi-omic analysis provided insights into the biological mechanisms underlying the metabolic benefits of meal timing matching an individual’s chronotype.

In terms of study limitations, the benefits of aligning the timing of TRE were only evaluated in individuals with a morning chronotype; future studies should test the same e-TRE vs. l-TRE vs. control paradigm in evening chronotype. Second, in e-TRE, the timing of meal intake deviated slightly from the intended schedule despite adherence to an 8h eating window. Future research should incorporate assessment of participants’ experiences and perceived challenges to clarify factors influencing adherence to the intended TRE schedule over a longer time period.^17,18,62,63^ Third, by study design, we included only people with BMI 25–34 kg/m^2^ and aged 25–50 years. Future studies are needed to evaluate the same interventions in individuals with higher BMI, postmenopausal women, or older populations. Fourth, participants could forgo or miss the regular recording of their food and drink consumption with the smartphone app. However, in previous studies, 3-4 days of recording seemed already reliable to assess macronutrient consumption,^74^ while 10-14 days were needed to additionally assess meal timing.^75^

In conclusion, our findings support the importance of matching eating-window timing with individual chronotype. For morning chronotype adults with overweight or obesity, aligning meals with chronotype through the early variant of TRE may enhance the metabolic benefits of TRE. Individuals at risk of excessive loss of lean mass, however, may require regular body composition monitoring with appropriate countermeasures in place. Notably, the two TRE variants offered complementary benefits: while e-TRE appears to enhance circadian glucose rhythmicity, l-TRE more effectively attenuates postprandial glucose excursions. In blood multi-omic analyses, the chronotype-aligned e-TRE induced a systemic signature with greater lipid remodeling and immune-cell metabolic adaptation.

## Methods

### Experimental study design

This three-arm, open-label, parallel-group RCT was conducted in a single center study at Geneva University Hospitals (HUG), Geneva, Switzerland. The study protocol was approved by the Ethics Committee of the Canton of Geneva, Switzerland (approval no. 2020-01439), and was registered on ClinicalTrials.gov registry (no. NCT04618133, first record on November 4, 2020) and the Swiss National Clinical Trial Portal (registry no. SNCTP000004143). All participants provided written informed consent prior to enrollment in the study. Data collection was conducted from January 2021 to October 2024. This study was carried out in accordance with the Declaration of Helsinki, the ICH Good Clinical Practices and the Swiss Human Research Act, with independent monitoring of the study.

Participants were recruited via flyers, posters, social media posts and online advertisements. We assessed 958 prospective individuals for eligibility criteria first (exhaustive list below). Participants were most commonly excluded for unstable weight (within ± 2 kg), out-of-range age and BMI, suspected eating disorders, neutral or evening chronotype, or existing adherence to a short eating window or intermittent fasting (**Figure S1**).

After checking all inclusion and exclusion criteria, participants underwent baseline assessment using questionnaires, wearable devices over two weeks, and fasting blood samples at 8–9 a.m. (detailed below). At visit 2, key inclusion criteria were verified objectively after two weeks (eating duration ≥ 12h, confident use of the smartphone food diary app MyFoodRepo, and stable body fat mass within ± 1 kg) to check their eligibility before randomization.

#### Inclusion criteria

##### Clinical criteria

- Men and premenopausal women
- Age 25–50 years
- BMI 25–34 kg/m^2^ (the estimation of body fat mass by BIA was validated for BMI ≤ 34 kg/m^2^)^76^
- Stable weight (maximum ± 2 kg) over the previous 3 months
- Stable body fat mass (maximum ± 1 kg of body fat mass, <u>key inclusion criterion</u>)
- Eating window ≥ 12h (self-reported and recorded between visits 1–2, <u>key inclusion</u> <u>criterion</u>)

##### Work and time-related criteria

- Morning chronotype (defined as Horne-Östberg Morningness-Eveningness Questionnaire^22^ (MEQ) scores ranging from ‘definitely morning’ (70-86 points) to ‘moderately morning’ (59-69 points))
- Daytime work at least 3 days per week over the previous month and planned during the study
- Study-related criteria
- Able to give informed consent and follow the study procedures during the study
- Confident use of a smartphone and able to take regular pictures of food and drinks with the MyFoodRepo smartphone app (<u>key inclusion criterion</u>)

#### Exclusion criteria

##### Clinical criteria

- Pregnant and breastfeeding women, plans for maternity during the study
- On a diet, intermittent fasting, in a weight management program over the previous 3 months or planned during the study
- Eating disorder(s) (screening with the Binge Eating Severity^77,78^ score ≥ 17) or prior bariatric surgery
- Diabetes with hypoglycemic drug(s)
- Major illness/fever over the previous month
- Active major cardiovascular, respiratory, liver, gastrointestinal, renal, neurological or endocrine disorders
- Coagulation disorder, on anticoagulant drug, skin disorder affecting wound healing
- Active cancer and/or oncologic treatment over the previous 12 months
- Major sleep disorder (including untreated sleep apnea syndrome), major mental illness
- Consumption of > 7 standard units of alcohol per week for women and > 14 standard units of alcohol per week for men

##### Work and time-related criteria

- Shift work, such as evening shifts or nights shifts over the previous month or planned during the study
- Travel/trip to a different time zone (≥ 2-hour time difference) over the previous month or planned during the study

##### Study-related criteria and other interventions

- Enrolled in another interventional clinical trial (medication, surgery, medical device) over the previous month or planned during the study
- Regular medications over the previous month that could affect the study endpoints (e.g. centrally acting, medications affecting gut absorption, transit or weight, hypoglycemic drug, hormonal treatment)

### Randomization and allocated intervention

Following the baseline measurements over two weeks (**Figure 1A**), participants were randomized with an allocation ratio of 1:1:1 to e-TRE (eating 06:00h–14:00h) vs. l-TRE (eating 12:00h–20:00h) vs. active control (three meals per day) for 12 weeks (**Figure 1B**, **Figure S1**). The randomization was performed on the REDCap platform, with a random allocation sequence stratified by sex. They were permitted one day per week without restrictions on meal timing, allowing greater flexibility for social and family activities. We followed each participant during the 12-week intervention phase with monthly phone calls to assess logging adherence with the MyFoodRepo smartphone app and the allocated intervention. Data collection was repeated over the last 2 weeks of the intervention phase (**Figure 1A**), totaling 6 visits (4 in person + 2 phone calls) per participant.

After randomization (**Figure S1**), two participants withdrew from the study (difficulty following the intervention, lack of motivation) and one was randomized by error (did not actually meet all eligibility criteria, which was discovered on the day following randomization). Recruitment continued until the target number of at least 17 participants per intervention group was reached to ensure statistical power. One participant allocated to e-TRE switched to l-TRE for socio-familial reasons and was analysed according to their original group assignment in the primary intention-to-treat analysis, and according to the actual intervention in the secondary per protocol analysis.

The power calculation was based on a literature review and assessment of clinical significance, based on known literature at the beginning of the study:

- Comparison 1: With an estimated mean loss of body fat mass of 0 ± SD 1 kg in the active control arm and an estimated mean loss of body fat mass of 2 ± 2 kg in the early TRE arm, 33 participants (i.e. 11 in each arm) were necessary to reach an 80% power (1-β) and two-sided significance level of p < 0.05 (α).
- Comparison 2: With an estimated mean loss of body fat mass of 0 ± 1 kg in the active control arm and an estimated mean loss of body fat mass of 1 ± 2 kg in the late TRE arm, 51 participants (i.e. 17 in each arm) were necessary to reach an 80% power (1-β) and two-sided significance level of p < 0.05 (α).
- Combining comparisons 1+2: To have sufficient power to detect a difference in the 2 planned comparisons, we kept the highest sample size estimation (i.e. hypothesis 2: 51 participants, 17 in each arm). The target sample size was therefore 51 participants who have completed the study intervention.

### Questionnaires

Clinical and questionnaire data were collected using the REDCap electronic data capture platform^79,80^ hosted at Geneva University Hospitals, Geneva, Switzerland. We recorded age, sex, demographics, medical history, lifestyle habits, personal experience and compliance with the intervention.

The chronotype was assessed using the Horne-Östberg MEQ,^22^ with scores ranging from ‘definitely morning’ (70-86 points), ‘moderately morning’ (59-69 points), ‘neutral’ (42-58 points), ‘moderately evening’ (31-41 points), to ‘definitely evening’ (16-30 points). By study design, only those with definitely or moderately morning chronotypes were included (see criteria above).

We assessed the self-reported sleep duration over the 4 previous weeks using the Pittsburgh Sleep Quality Index.^81^ Self-reported physical activity levels over the 4 previous weeks were assessed using the short form of the International Physical Activity Questionnaire,^82^ with a total score and three subscores for low, moderate and high physical activity levels.

### Clinical measurements and fasting blood tests

Weight was measured in light clothing with an electronic scale calibrated yearly, and the BMI was calculated as the weight in kilograms divided by the height in meters squared (kg/m^2^). Waist and hip circumferences and waist-to-hip ratio (WHR) were assessed according to the WHO standard procedure (ISBN 978-92-4-150149-1). Blood pressure was measured three times with a calibrated monitor and an appropriately sized arm cuff, after 5 minutes of rest in the sitting position, and the last two values were averaged.

Fasting plasma glucose, insulin, glycated hemoglobin (HbA1c), and lipid profile (total, HDL and LDL cholesterol, triglycerides) were measured in the fasting state at 8–9 a.m using the routine assays at the clinical lab of Geneva University Hospitals, Geneva, Switzerland. Insulin resistance was assessed with the HOMA-IR, based on the fasting plasma glucose and insulin.^83,84^

### Body composition (primary outcome)

Body fat mass (the primary outcome), lean mass and bone mass were assessed by dual-energy X-ray absorptiometry^85,86^ (DXA) using the Hologic Discovery A (Hologic, Waltham, MA, USA) at visits 2 and 6. Soft tissue can further be divided into body fat mass and lean mass as they have different attenuation characteristics. The visceral fat mass, also known as visceral adipose tissue, was computed by subtracting subcutaneous adipose tissue from the total android fat mass.^87,88^

Fat mass and fat-free mass were estimated from electric resistance and reactance measured by BIA using the Nutriguard-M^®^ device (Data Input, Germany). Skin adhesive electrodes were placed on the right hand and foot, while the subject was lying on their back. An alternating electrical current of 50 kHz and 0.8 mA was applied. The measured resistance and reactance were used to estimate fat-free mass by a formula developed in Geneva and validated against DXA.^76^ Fat mass was calculated by subtracting fat-free mass to body weight. Fat mass index and fat-free mass index were calculated by dividing them by the height squared (kg/m^2^).

### Resting energy expenditure

Resting energy expenditure (REE) was measured by indirect calorimetry, at rest in the supine position, after an overnight fast just before the fasting blood tests (and the optional skin biopsy). After calibration of O_2_ and CO_2_, and adjustment for bias flow, the Q-NRG^®^ (Cosmed, Pavone, Italy) device measures VO_2_ through a paramagnetic analyzer and VCO_2_ through infrared absorption. These two parameters allow the calculation of the resting energy expenditure by the Weir equation:^89^

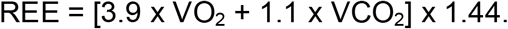

### Actigraphy

Actigraphy was assessed using a triaxial accelerometer (GENEActiv, Activinsights Ltd, United Kingdom) worn on the non-dominant wrist and recording with a sampling frequency of 50 Hz. Data on daily activity and sleep/wake cycles were analysed with the GGIR package in R (v4.0.0),^90^ based on validated intensity cutoffs with the following published values:^91^ light intensity physical activity (45.8-93.2 mg), moderate intensity physical activity (93.2-418.3 mg), and vigorous intensity physical activity (> 418.3 mg). Other GGIR analysis parameters included an epoch length of 5 seconds, days defined from waking-up to waking-up, a valid day included ≥ 16 hours of data. From the GGIR sleep and physical activity analysis output, we extracted the sleep onset, wakeup time, sleep duration, light, moderate and vigorous physical activity.

### Smartphone food diary app

The smartphone food diary app MyFoodRepo was developed at the Digital Epidemiology Lab, EPFL, Switzerland, and records consumed food and drink along with pictures, barcodes of packaged items, or free text descriptions.^44^ Based on pictures and a chat function, each entry is then validated by trained dieticians helped by computer vision and constantly updated nutritional composition table, and the data output contains a list of all consumed food and drink items, and timestamped calorie and macronutrient intake. The accuracy of this food diary app has been validated against reference values from weighed food diaries.^43,45^ A minimum number of 2-3 days were required to obtain reliable estimates of dietary intake in a large digital cohort.^74^

The eating window was defined as the time duration between the first and the last calorie-containing meal. The mean consumption rate per hour was calculated by using a kernel density estimate (with a von Mises distribution, κ = 30) with grid points spanning from 0 to 24 hours every 15 minutes, which was then normalized such that the total area under the curve equaled the total daily consumption.

### Continuous Glucose Monitors (CGM)

CGM data was recorded with an Abbott FreeStyle Libre^®^ Pro device (Abbott Diabetes), which records interstitial glucose every 15 mins. During the initial period of recording, the CGM data can occasionally spend the majority of the time at the device minimal value of 2.2 mmol/L due to the transient effects of microtrauma, local pressure leading to lower blood flow and device calibration. In this case, we removed the initial data until the device exited this floor state. As a preliminary step of glucose dynamics modelling (see below), abnormally low glucose values (that are typical of the early recording hours) were also corrected by removing long timescale trends within the time series. CGM metrics related to glycemic control were extracted from the CGM data using the R package rGV,^92^ and we focused on the mean glucose level, coefficient of variation (CV = SD/mean, expressed in %), and the mean amplitude of glycemic excursions (MAGE, expressed in mmol/L).^93^ To calculate the moving average 24-hour profile, we defined grid points spanning from 0 to 24 hours every 15 minutes and computed the average values over a 2-hour window centered on each grid point.

### Glucose dynamics – Modeling ingestion events and glucose data

We combined the ingestion events with CGM data based on a recently published computational framework^46^ (**Figure 5A**). The food and drink consumption entries recorded with the MyFoodRepo smartphone app are used to model postprandial interstitial glucose increases, and both the height of the glucose excursions (termed “response heights”) and the time needed for glucose to return to baseline (termed “response half-life”) are inferred for each individual at baseline and follow-up, independently. The model also includes an underlying 24-hour cosinor function to describe 24-hour oscillations in baseline (i.e. irrespective of meal intake) glucose levels, where the cosinor function assumes baseline glucose levels follow a sinusoidal pattern over the course of a day. The 24-hour cosinor function is described by the three parameters ‘baseline’, ‘peak-to-trough amplitude’ and ‘peak time’, which are similarly learned for each participant at baseline and follow-up, independently. The model was trained by minimizing the error between the model and the CGM data using a maximum *a posteriori* probability (MAP) estimate from 200 different starting conditions, and the settings and parameters of the original method were left unchanged.

### Missing data

The MyFoodRepo data on calorie intake, meals and beverages were assessed for all participants, except one with only 18 records over 2 weeks. No calorie filtering was performed. The CGM metrics were calculated when at least 48 hours of data were recorded. In certain cases, the CGM device fell off during the recording period and was replaced as soon as possible. The MyFoodRepo data was analyzed alongside the CGM data when both MyFoodRepo data and CGM data were concomitant and not missing. GENEActiv data was used for subsequent GGIR analysis when at least 6 days of data were available, and we also filtered participants with more than 20% missing data.

### Primary skin fibroblast culture, *in vitro* synchronization and real-time bioluminescence recording

Cutaneous biopsies were collected in the fasting state (8–9 a.m.) at baseline (visit 2) and after 12 weeks (visit 6) from a subset of participants (n = 5 per intervention group). Biopsies were processed as previously described.^58,94^ In short, skin fibroblasts were cultured in DMEM 1g/L D-glucose, with pyruvate and GlutaMAX supplemented medium (Gibco, Paisley, Scotland, UK), enriched in 15% fetal bovine serum (Gibco, Paisley, Scotland, UK), 1 % penicillin/streptomycin (Gibco, Paisley, Scotland, UK), 0.5 % gentamycin (Sigma Aldrich, Saint Louis, USA) and 0.2% amphotericin B (Gibco, Paisley, Scotland, UK). Cell cultures were transduced with Bmal1-luciferase lentivectors with a MOI = 3 and synchronized with 100 nM of dexamethasone (Sigma Aldrich, Saint Louis, USA) for 30 minutes 3-5 days post-transduction and subjected subsequently to real-time recording of Bmal1-luciferase bioluminescence for a minimum of 100 hours in the presence of luciferin. Technical triplicates were processed for each cell line.

Generated bioluminescence profiles were normalized to the 24-hour moving average (detrended).^95^ Circadian parameters were calculated based on detrended data using cosinor-lite (https://github.com/Nick-E-P/cosinor-lite/tree/main/src/cosinor_lite) that uses a cosinor analysis of the 12-104h data range. The circadian period was estimated to be within a 20–28h window.

### Statistical analyses

Continuous and normally distributed variables are reported as mean ± SD and compared with paired *t* tests. Non-normally distributed continuous variables are reported as median (interquartile range, IQR) and compared with the Mann-Whitney U test (i.e. Wilcoxon signed-rank test). Time measures are expressed as clock time (hh:mm), while durations are expressed as decimal hours (h). Categorical variables are presented as the number of participants (% in group) and compared using the chi-squared test. Paired statistical tests were performed using the ‘pingouin’ package (v0.5.3) within Python.

We used analysis of covariance (ANCOVA) to perform statistical comparisons between groups, with the following regression equation for each variable analyzed:^96^

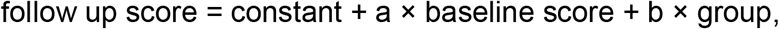

where group is a binary variable with the following codings for the three comparisons: early TRE vs active control (early TRE = 1, active control = 0), late TRE vs active control (late TRE = 1, active control = 0), early TRE vs late TRE (early TRE = 1, late TRE = 0). The analysis was performed using the ‘statsmodels’ (v0.13.0) package within Python. To account for multiple comparisons, p-values for secondary and exploratory outcomes were adjusted using the Benjamini-Hochberg procedure, with both adjusted and unadjusted p-values reported in the corresponding tables.

### Plasma, serum, and monocyte pre-processing

Serum samples were collected in the fasting state (8–9 a.m.) at baseline (visit 2) and after 12 weeks (visit 6) for metabolomic and lipidomic profiling. All mass spectrometry and sequencing analyses were conducted at the same institutional core facilities using established workflows consistent with our prior studies.

For metabolomics, 15 µL of serum was spiked with heavy isotope-labeled internal standards prior to extraction. Metabolites were extracted using a liquid-liquid extraction and protein precipitation with methanol. Analyses were performed on a 1290 Infinity II UHPLC system (Agilent Technologies) coupled to a 6470 triple quadrupole mass spectrometer operating in negative electrospray ionization mode. Chromatographic separation was achieved on a ZORBAX RRHD Extend-C18 column (2.1 × 150 mm, 1.8 μm) maintained at 40°C using a tributylamine/acetic acid-buffered gradient. Metabolites were detected using dynamic multiple reaction monitoring and quantified using internal standard calibration curves. Raw data was processed using PeakBot software.

For lipidomics, 20 µL of serum, previously spiked with heavy isotope-labeled internal standards, was extracted using methyl tert-butyl ether and methanol according to a modified Matyash protocol. Lipid separation was performed by reversed-phase chromatography on an Accucore C18 column (150 × 2 mm, 2.6 μm) maintained at 35°C, using a binary solvent system containing ammonium formate and formic acid. LC-MS analysis was carried out on a Vanquish UHPLC system coupled to an Orbitrap Fusion Lumos Tribrid mass spectrometer (Thermo Fisher Scientific), operated in both positive and negative ionization modes at a resolution of 120,000 over an *m/z* range of 250-1200. Lipid peak detection and quantification were performed using TraceFinder software.

For metabolomics and lipidomics, values below the limit of detection were set to missing, and features with more than 50% missing values across all samples were removed. Remaining missing or zero values were imputed with one fifth of the smallest positive value of that feature. Samples were then normalized by probabilistic quotient normalization (PQN) and log_2_-transformed as log_2_(x + 0.5 × the smallest positive PQN value). Only participants with both visits were kept.

Peripheral blood mononuclear cells were isolated using CPT tubes (BD Vacutainer CPT) followed by centrifugation at 1,800g. Cells were washed in PBS, cryopreserved in 10% DMSO-containing freezing medium, and stored at −80°C. CD14+ monocytes were enriched using magnetic bead separation (Miltenyi Biotec) and purified using an AutoMACS Pro separator. Total RNA was quantified by Qubit fluorometry and integrity assessed using Bioanalyzer/TapeStation systems (Agilent Technologies). Libraries were prepared from 200 ng total RNA using the Illumina Stranded mRNA Prep kit and sequenced on a NovaSeq 6000 platform (100-bp single-end reads) at the iGE3 Genomics Platform (University of Geneva). Reads were quality-controlled using FastQC, aligned to the human reference genome (hg38) using STAR, and gene-level counts were generated using HTSeq. RNA-seq counts were filtered, normalized (TMM) and converted to log-CPM using the ‘edgeR’ (v4.8.2) package within R.

### Internal circadian time inference from blood monocyte RNA-seq

Internal circadian time was estimated from the monocyte RNA-seq with two published algorithms, TimeMachine^60^ and ZeitZeiger.^61^ Both were trained locally on the reference dataset distributed with TimeMachine (170 training samples with known circadian time) and restricted to the cycling genes of the TimeMachine gene panel detected in our data (35 of 37; NELL2 and EPHX2 were absent). Predictions were evaluated on the 596 held-out samples of the reference dataset, and we applied the same criterion to every method: a median absolute error ≤ 3h. TimeMachine predicts internal time from the pairwise log_2_ ratios of the panel genes within each sample (595 ratios per sample), which removes platform and batch offsets. The elastic-net model (α = 0.2 and λ = 0.31, published settings) returns a predicted amplitude and samples with amplitude below 0.5 were considered unreliable (0 out of 50). In ZeitZeiger, expression was z-scored within each sample across the panel genes for the same reason. The model fits a periodic spline to each gene, extracts sparse principal components of the time-dependent signal, and predicts time by maximum likelihood. The sparsity parameter (3) and the number of components (2) were selected on the held-out sample. Both models predicted circadian time in held-out samples with a median absolute error of 2.2h and 2.7h, respectively. Agreement between the methods was quantified by Spearman correlation of per-subject shifts. Shifts from baseline to follow-up were tested against zero within each group (one-sample t test), and groups were compared by ANCOVA. Analyses were performed using ‘glmnet’ (v5.1) and ‘zeitzeiger’ (v2.1.3) packages, and TimeMachine was run using the authors’ script (github.com/pepperhuang/TimeMachine; accessed 14 October 2026) within R.

### Single-omics block analysis

Features were tested with limma-voom workflow using the model ∼ Sex + Group × Time, accounting for repeated measures by blocking for participants. For transcripts, the model was run first without blocking to estimate the correlation, then with the block and consensus correlation. Contrasts were defined as the within-group changes (baseline to follow-up) and as the between groups change (difference in change). Moderated statistics came from robust empirical Bayes. P-values were adjusted by Benjamini-Hochberg within each contrast and omics layer. Volcano plots show FDR against log_2_ fold-change and only q < 0.05 were colored.

To interpret the feature-level results at a higher level, we tested for coordinated changes in sets of functionally related features (metabolic pathways, lipid classes, transcriptional programs) instead of single features. Features were ranked by the moderated t-statistic of each limma-voom contrast, so that no fold-change or significance threshold had to be applied to individual features. The method differed by layer. Metabolic pathways were mapped to Kyoto encyclopedia of genes and genomes (KEGG) compound IDs. Pre-ranked set enrichment was run on KEGG pathways (10–500 members; 10,000 permutations). Lipid set enrichment analysis^97^ was restricted to lipid class (at least 4 members; 10,000 permutations). For transcripts, we used sample-level gene-set enrichment scores (GSVA) instead of a ranked list, for consistency with the multi-block analysis and because gene-level changes are small and correlated within pathways, and rank-based enrichment permutes genes as if they were independent. Scoring first and testing the scores in the repeated-measures limma-voom model moves the inference to the participants. Molecular signatures database (MSigDB) hallmark activity was scored per sample and visit (≥ 10 genes with mean log_2_-CPM ≥ 0 and covering ≥ 50% of the hallmark set). The scores were analyzed with limma-voom using the model ∼ 0 + Sex + Group x Time, with participants as blocking factor and with robust empirical Bayes.

All block set enrichments were corrected with Benjamini-Hochberg across sets within each contrast. The analyses were performed using the ‘limma’ (v3.66.0), ‘fgsea’ (v1.36.2, for metabolomics), ‘lipidr’ (v2.24.0, for lipidomics) and the ‘GSVA’ (v2.4.9, for transcriptomics) packages within R.

### Multi-block integrative modelling

For each participant, the change Δ *= follow-up – baseline* was computed on log_2_ metabolite and lipid values. For transcripts, Δ was computed on hallmark GSVA scores as described above. This gives a transcriptomic block of a size comparable to the other layers, avoids an arbitrary choice of a variable-gene subset, and summarizes small, correlated gene-level changes into interpretable biological programs. Only participants that have data in all three omic blocks were included (total n = 25; e-TRE n = 8, l-TRE n = 8, active control n = 9). The final blocks were 93 metabolites, 225 lipids and 43 hallmark scores.

We followed the approach by Grismer,^98^ which combines multiple factor analysis (MFA) with permutational multivariate analysis of variance (PERMANOVA). MFA is a multi-block extension of principal component analysis. It summarizes the variation shared across several sets of variables measured on the same participants in a small number of dimensions. It is unsupervised, so the dimensions are defined without reference to the intervention groups. MFA was run on the matrix of participants by within-participant changes in each feature. Each omics layer was one block of quantitative variables, and each variable was centered and scaled to unit variance. MFA weighs each group by the inverse of the first eigenvalue of its own PCA, so no block dominates through size or variance. The first ten dimensions, specified in advance, were retained for all downstream tests.

PERMANOVA then tests whether the intervention groups differ in their multivariate location (centroid) in the space defined by these dimensions. It is a permutation-based test that makes no distributional assumptions, which suits small groups. Euclidean distances between participants were computed on the first ten MFA dimensions and tested with adonis2() (group as the factor; 9999 permutations; seed = 2025). We tested the omnibus effect and the three pairwise contrasts (e-TRE vs. active control, l-TRE vs. active control, e-TRE vs. l-TRE). R^2^ is reported as the effect size. Homogeneity of multivariate dispersions was assessed with a permutational analysis of multivariate dispersions (PERMDISP; betadisper() followed by a permutation test with 9999 permutations) on the same Euclidean distances. The analysis was performed using the ‘FactoMineR’ (v2.18) and ‘vegan’ (v2.7.6) packages within R.

## Data Availability

Requests for further information and resources should be directed to and will be fulfilled by the lead contact, Tinh-Hai Collet. Metabolomics and lipidomics data supporting the findings of this study will be made available in Data S3 soon after submission. Raw RNA-seq data will be deposited in GEO soon after first submission. Any additional information required to reanalyze the data reported in this paper is available from the lead contact upon request.

## RESOURCE AVAILABILITY

### Lead contact

Requests for further information and resources should be directed to and will be fulfilled by the lead contact, Tinh-Hai Collet.

### Materials availability

This study did not generate new, unique reagents.

### Data and code availability

Metabolomics and lipidomics data supporting the findings of this study will be made available in Data S3 soon after submission. Raw RNA-seq data will be deposited in GEO soon after first submission. Any additional information required to reanalyze the data reported in this paper is available from the lead contact upon request.

## ACKNOWLEDGMENTS

The authors wish to thank all participants, the clinical and research team at the Clinical Research Center, Geneva University Hospitals and Faculty of Medicine, University of Geneva, the laboratory team at the Geneva University Hospitals Sérothèque (Biobank), the team of the Digital Epidemiology laboratory, EPFL, and the annotators of recorded food and drink pictures via the MyFoodRepo smartphone app. The authors gratefully acknowledge the Metabolomics team from the Molecular Discovery Platform, CeMM Research Center for Molecular Medicine of the Austrian Academy of Sciences, for the metabolomics and lipidomics analyses, as well as Miriam Abele, Lenka Novotna, and Nara Marella for technical support, data acquisition, and assistance in these analyses. The authors have not used any large language models to generate the text, tables and figures of this manuscript.

## AUTHOR CONTRIBUTIONS (following the CRediT taxonomy)

Study conception: J.M., J.J.P., C.D. and T.H.C.; Methodology: N.E.P., L.M., J.M., F.S., J.J.P., C.D. and T.H.C.; Computation and software: N.E.P., L.M., A.D.B., M.S. and T.H.C.; Formal analysis: N.E.P., L.M., C.D. and T.H.C.; Investigation – data collection: J.M., F.S., C.J., J.S.A., A.H., O.F., S.B., D.G. and T.H.C.; Investigation – data curation: N.E.P., F.C.J., L.M., J.M., F.S., C.J., A.H., and S.B.; Writing – initial draft: N.E.P., F.C.J., L.M. and T.H.C.; Writing – review and editing: all authors; Writing – visualization and data presentation: N.E.P., L.M., A.D.B. and T.H.C.; Supervision: J.J.P., C.D. and T.H.C.; Project administration: T.H.C.; Funding acquisition: J.J.P., C.D. and T.H.C.

## SUPPLEMENTAL INFORMATION

Supplementary Results, Figures S1-S8, Tables S1-S5.

---

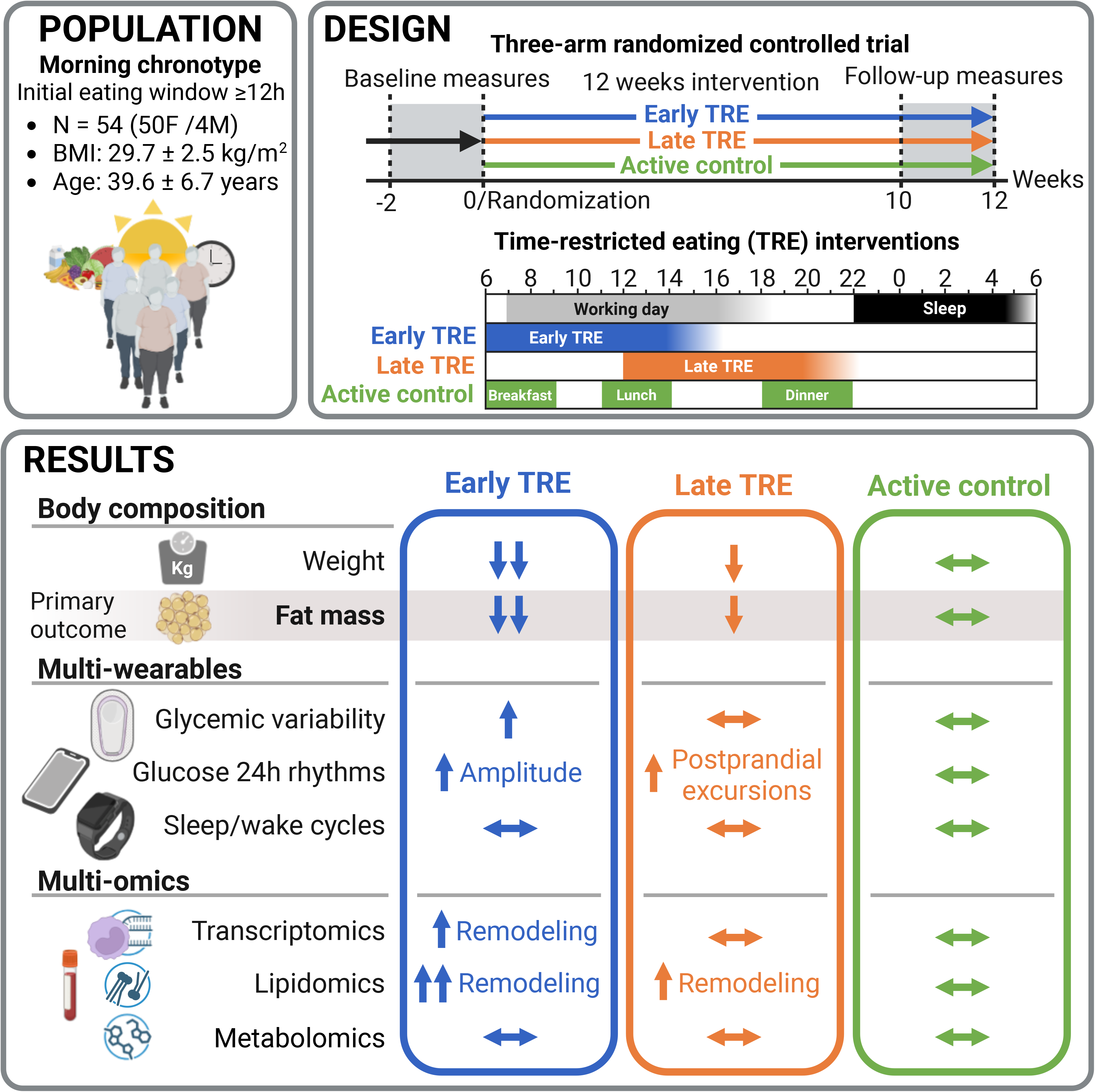

