## Supplemental Information for "Early Time-Restricted Eating Reduces Fat Mass In Morning Chronotype Adults With Overweight/Obesity"

#### Table of contents

Supplementary Results

Supplementary Figures S1–S8

Supplementary Tables S1–S5

### SUPPLEMENTARY RESULTS

**Early TRE led to reduced fasting plasma glucose, insulin resistance and sleep duration** *(related to the Results section of the same name)*

We assessed changes in metabolic health by clinical measurements, indirect calorimetry, and metabolic blood profile in the fasting state at 8–9 a.m. (**Table S3**). There was a decrease of fasting plasma glucose and insulin resistance (HOMA-IR) with early time-restricted eating (TRE) compared to the active control intervention (both p = 0.03), but these differences did not remain statistically significant after correction for multiple testing. Overall, there were no statistical changes in blood pressure, HbA1c, or lipid profile after the 12-week long intervention within and between groups (**Table S3**).

The resting energy expenditure (REE) decreased with early TRE (median –56 kcal/d, IQR –139 to –11, p = 0.02), but there were no statistical differences across groups (**Table S3**). In addition, there was a slight decrease of the respiratory quotient after early TRE compared to late TRE (–0.04, 95%CI –0.08 to –0.01, unadjusted p = 0.007, after adjustment for multiple testing p = 0.17), suggesting a shift towards greater fat oxidation. This difference may be explained by the longer interval between the last calorie intake (end of the eating window) and the REE measurement the following morning in the early TRE group (15:50h ± SD 01:25h) compared to the late TRE (19:50h ± 00:15h) or active control groups (20:20h ± 00:40h, **Figure 1D**).

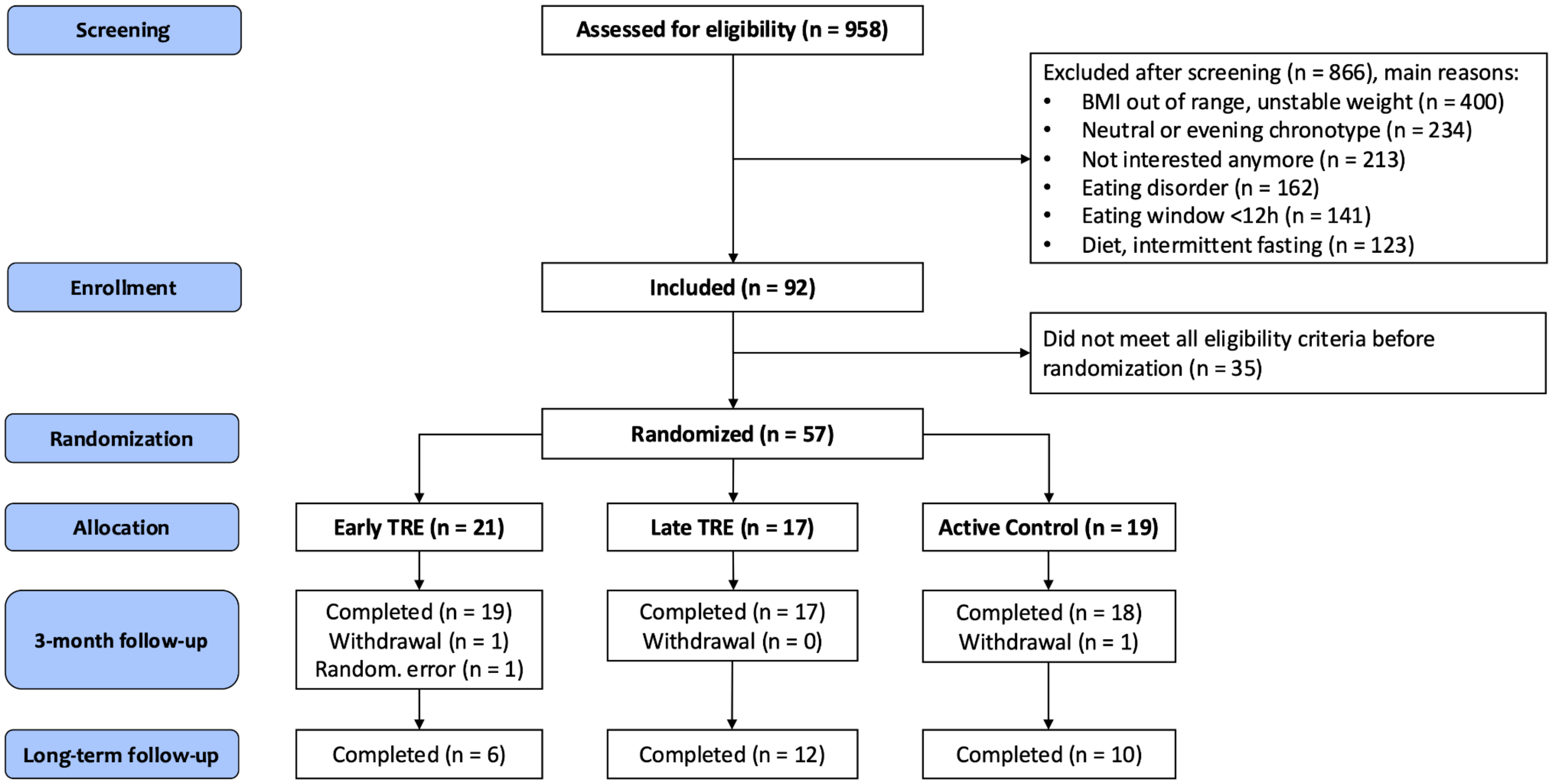


**Figure S1**. Flowchart of the study participants from screening, assessment of inclusion/exclusion criteria, enrollment and randomization, and completion of follow-up. The number of excluded individuals and the reasons for exclusion are detailed in the text boxes and in the Methods. One participant was randomized to early TRE intervention by error, while they did not meet all eligibility criteria (eating window < 12 hours, an exclusion criteria). This randomization error was identified the following day and this participant was excluded from the 12-week intervention phase; consequently, their data were not included in the follow-up analyses. *Related to Table 1, Figure 1.* Abbreviation: BMI, body mass index; TRE, time-restricted eating.

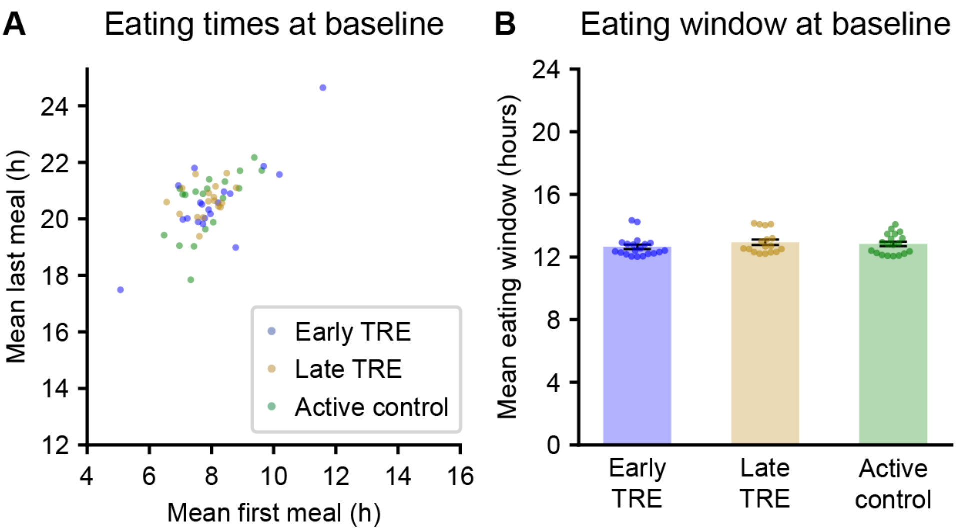


**Figure S2**. Chrononutrition behaviour of study participants at baseline, i.e. before randomization to early TRE, late TRE, or active control. (A) The mean timing of the first and last calorie intake for study participants during the baseline phase, and (B) the mean duration of the eating window (time elapsed between first and last calorie intake) during the baseline period, as measured with the smartphone food diary app MyFoodRepo. *Related to Figure 1D-E.* Abbreviations: h, hours; TRE, time-restricted eating.

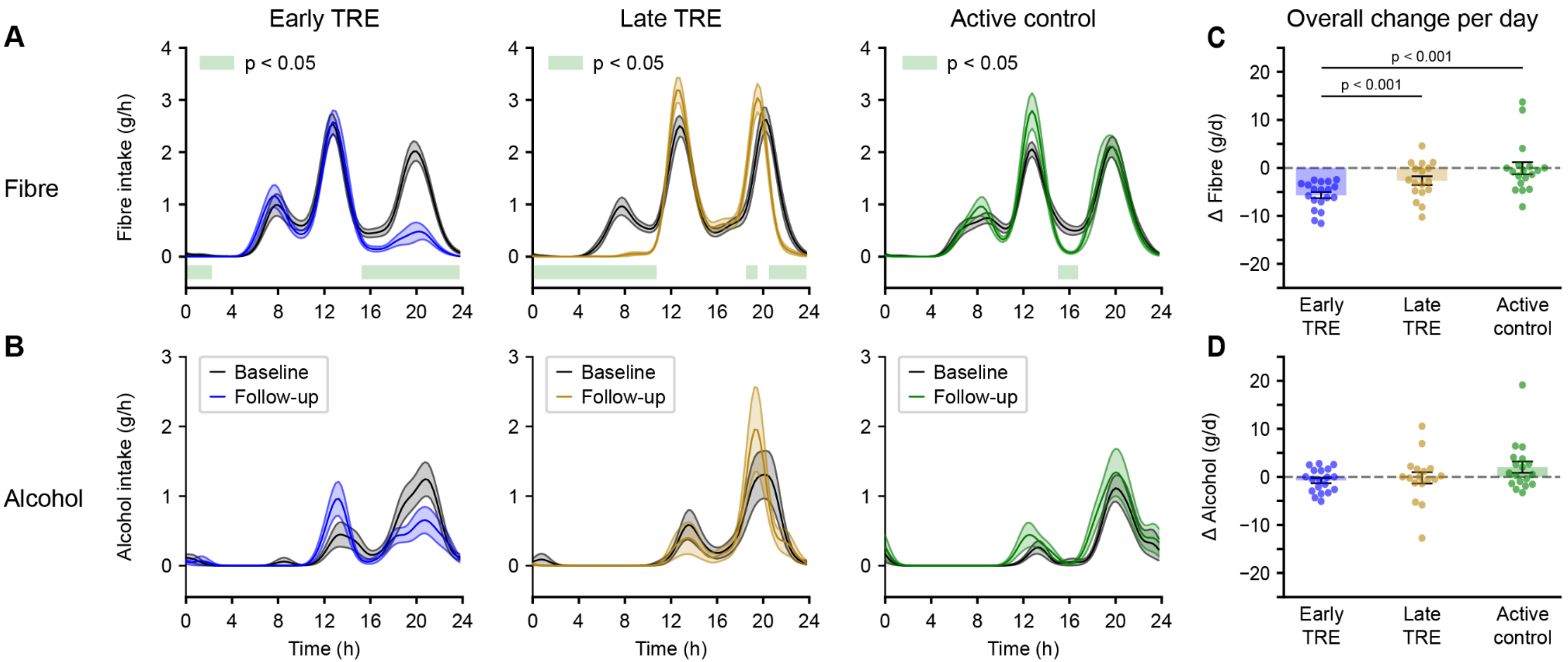


**Figure S3**. Changes in the timing and total intake of fibre and alcohol. (A-B) The mean consumption rate per hour for the early TRE (blue), late TRE (orange) and the active control group (green) for fibre and alcohol at baseline (black line) and follow-up periods (colored line). The shaded area represents the mean ± SEM, and the green bar at the bottom represents significant differences from baseline, as assessed from $p < 0.05$ in a Wilcoxon non-parametric test, adjusted for multiple testing with the Benjamini-Hochberg procedure. (C-D) Individual average change in daily intake for early TRE (blue), late TRE (orange) and active control (green) for fibre and alcohol, respectively. *Related to Figure 3, Table S4*. Abbreviations: d, day; h, hours; TRE, time-restricted eating.

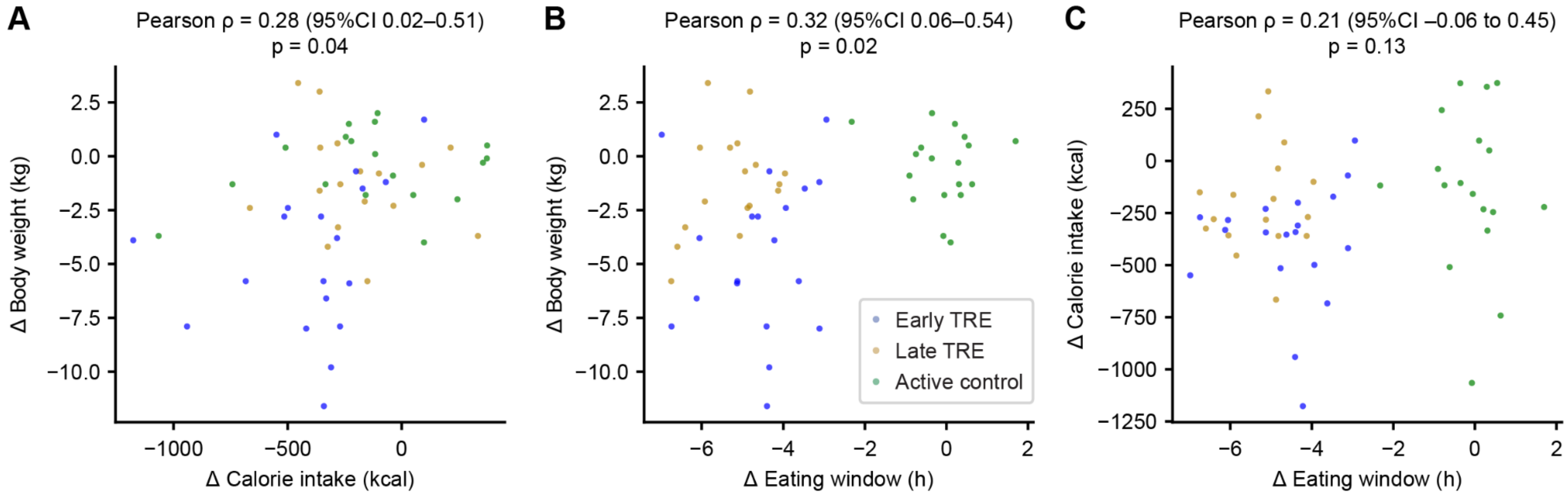


**Figure S4**. Change in body weight as a function of (A) change in calorie intake and (B) eating window, comparing post- and pre-intervention for the early TRE (blue), late TRE (orange) and active control (green). (C) Change in calorie intake as a function of the change in eating window, comparing post- and pre-intervention phases in each group. *Related to Figure 3*. Abbreviations: CI, confidence interval; h, hours; TRE, time-restricted eating

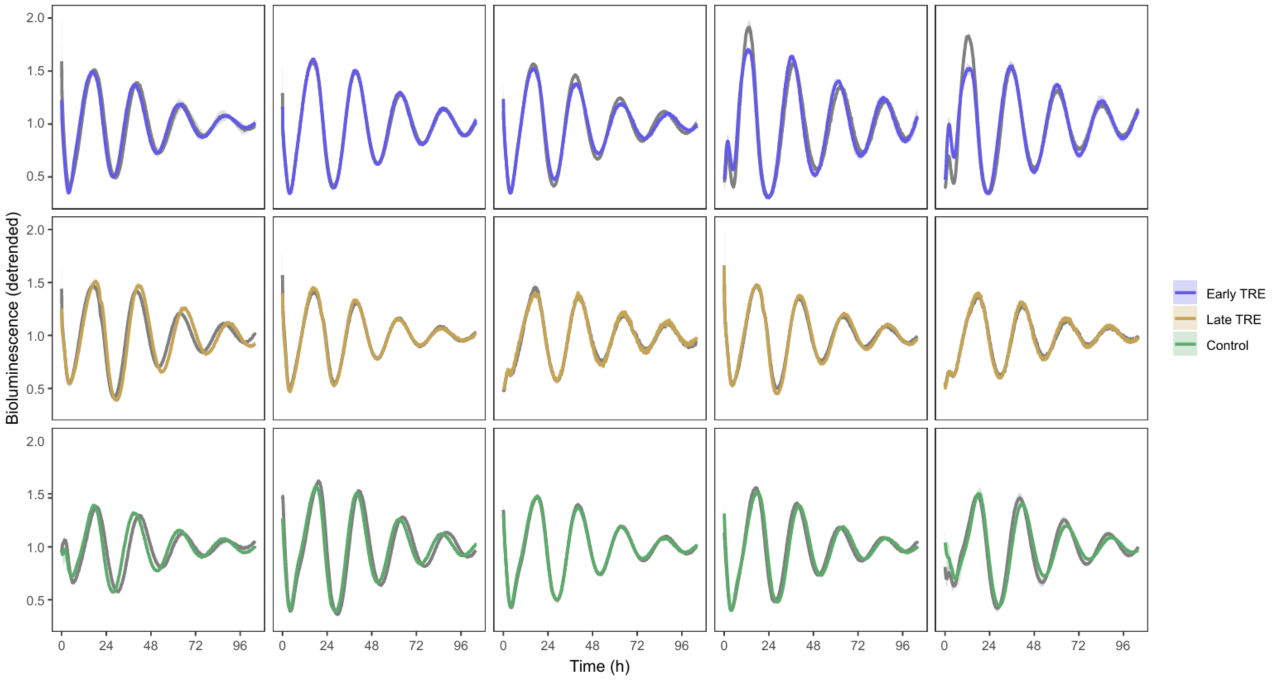


**Figure S5**. Individual molecular clock oscillations of primary skin fibroblasts measured by *Bmal1*-luciferase reporter assay. Averaged paired profiles mean ± SD (three technical replicates per subject at baseline and post-intervention) detrended values. Baseline fibroblasts oscillations are shown in grey and post-intervention oscillations are shown in color. Abbreviations: TRE, time-restricted eating.

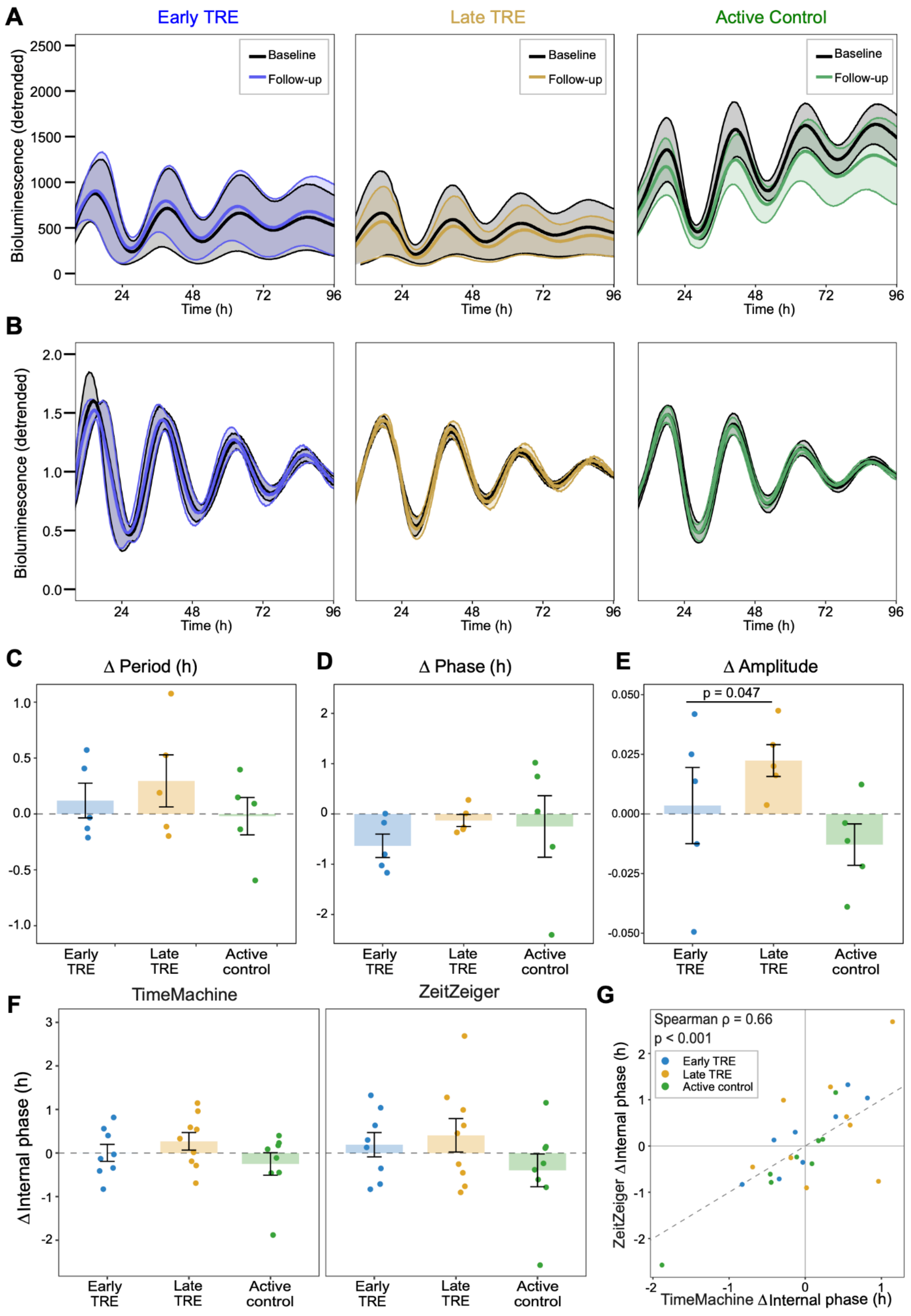


**Figure S6. Molecular clock properties of primary skin fibroblasts and monocyte-derived internal circadian phase are not affected by TRE.** (A–B) Averaged paired profiles ± SEM (n = 5 per regimen) with raw (A) and detrended (B) values. (C–E) Changes from pre- to post-intervention of the circadian parameters extracted from a cosine fit for the circadian period (C), phase (D), and amplitude (E). (F) Change in internal circadian phase from pre- to post-intervention (e-TRE, n = 8; l-TRE, n = 9; active control, n = 8) estimated from monocyte RNA-seq with TimeMachine (left) and ZeitZeiger (right). Baseline to follow-up within-group changes with TimeMachine (e-TRE 0.00h, $p = 0.98$; l-TRE +0.27h, $p = 0.54$; active control 0.25h, $p = 0.54$) and ZeitZeiger (e-TRE 0.19h, $p = 0.52$; l-TRE +0.41h, $p = 0.49$; Active control –0.40h, $p = 0.49$). Between-group changes with TimeMachine (e-TRE vs. active control -0.17h, $p = 0.84$; l-TRE vs. active control –0.14h, $p = 1$; e-TRE vs. l-TRE –0.10h, $p = 1$) and ZeitZeiger (e-TRE vs. active control -0.12h, $p = 1$; l-TRE vs. active control 0.54h, $p = 0.68$; e-TRE vs. l-TRE –0.44h, $p = 1$). (G) Agreement between the per-participant phase shifts estimated by TimeMachine and ZeitZeiger (Spearman $\rho = 0.66$, $p < 0.001$, $n = 25$). (C-F) Bars show the mean ± SEM and dots show individual participants. Pairwise group comparisons were calculated using ANCOVA and adjusted for multiple comparisons with Benjamini-Hocheberg. Abbreviations: ANCOVA, analysis of covariance; TRE, time-restricted eating.

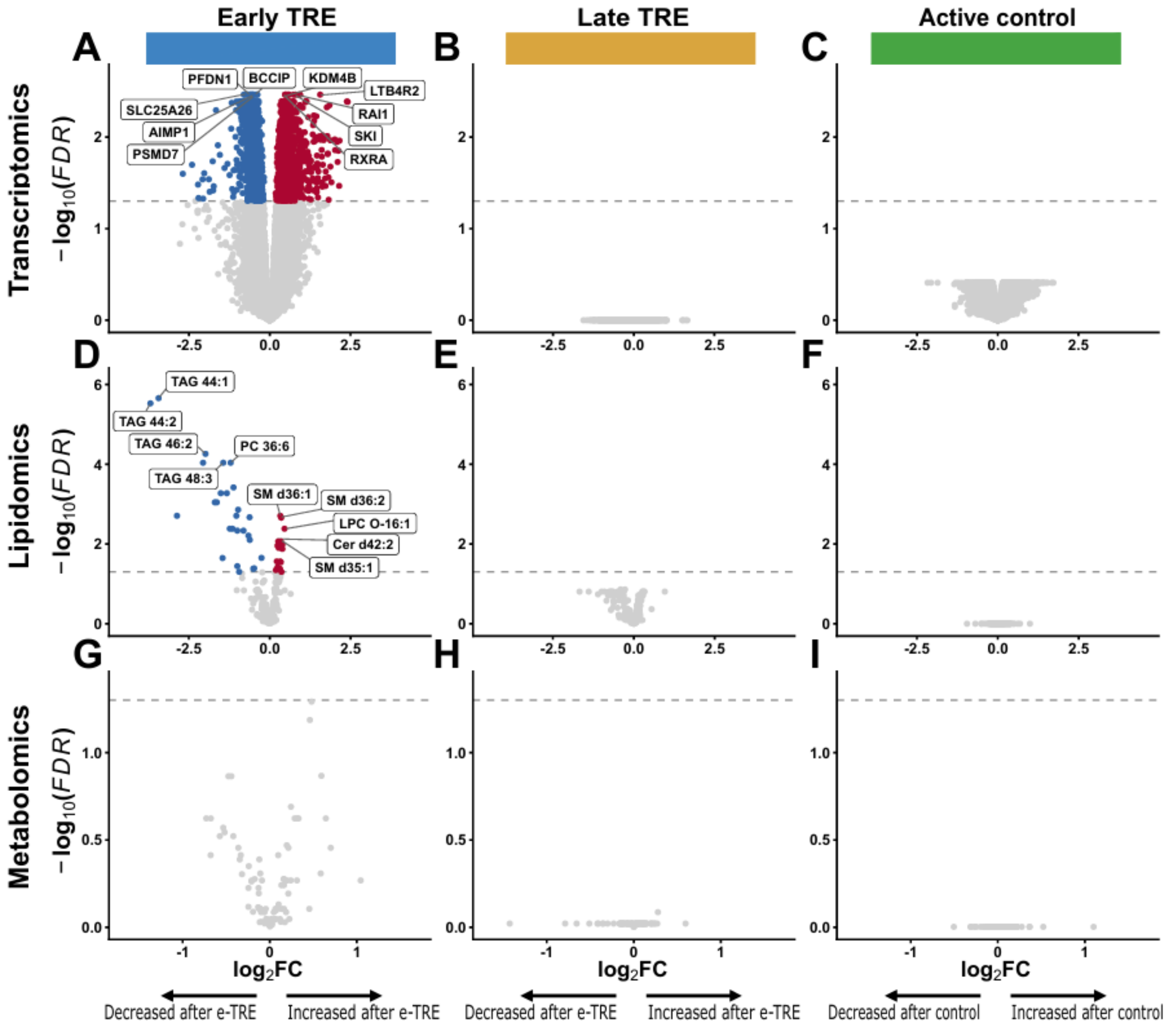


**Figure S7. In within-group comparisons, early TRE shows the most changes in blood multi-omics after intervention.** Volcano plots illustrate longitudinal changes from baseline to follow-up for early TRE (A, D, G), late TRE (B, E, H), and active control (C, F, I) groups. Data layers include monocyte transcripts (A-C), lipid species (D-F), and metabolites (G-I). Blue and red points denote features significantly decreased or increased (FDR < 0.05). Abbreviations: Cer, ceramides; FDR, False discovery rare; LPC-O, ether lysophosphatidylcholine; PC, phosphatidylcholines; SM, sphingomyelins; TAG, triacylglycerols; TRE, time-restricted eating.

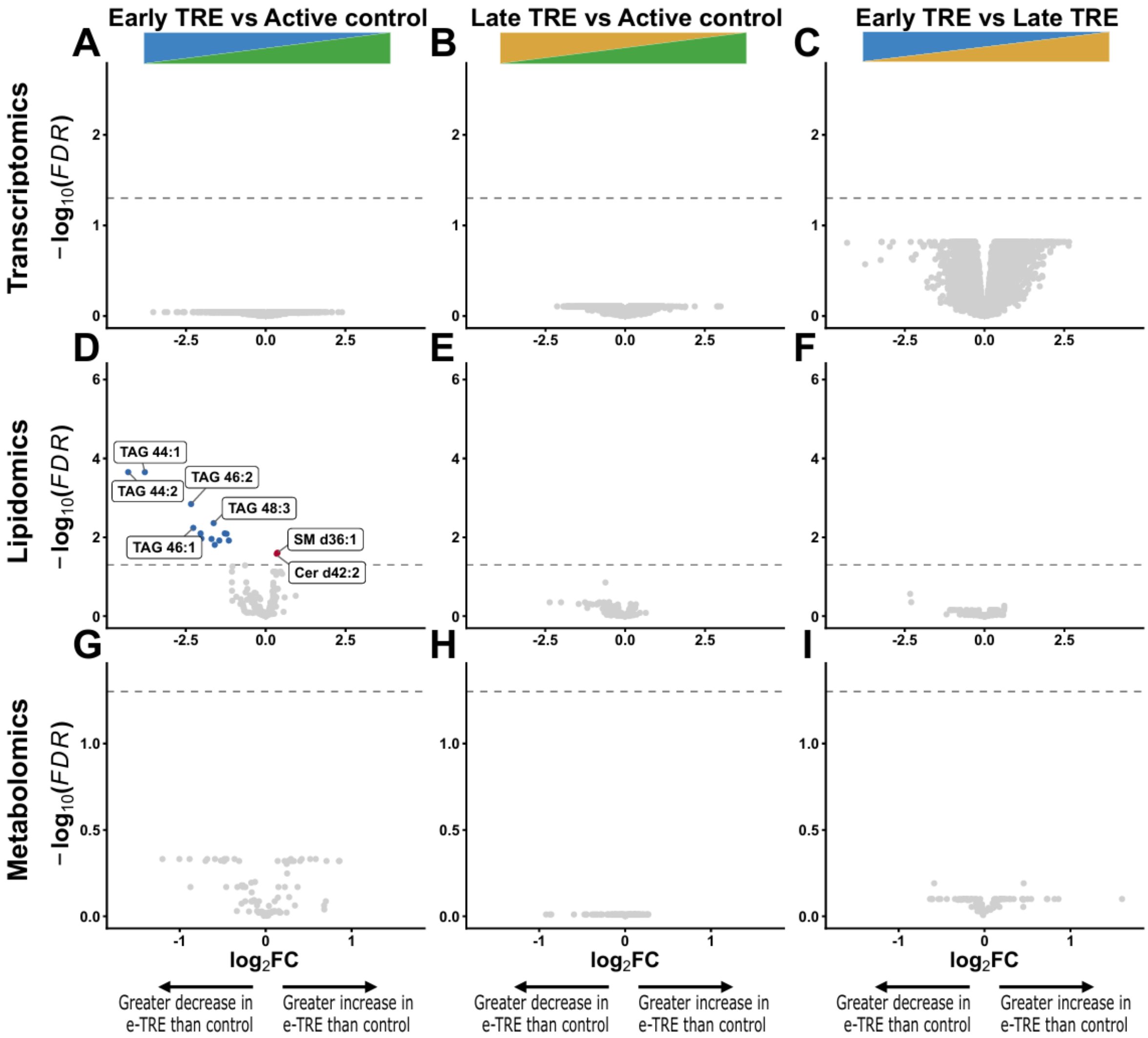


**Figure S8. In between-group comparisons, early TRE shows greater changes in blood multi-omics compared to late TRE and control.** Volcano plots illustrate the differences in changes between early TRE and active control groups (A, D, G), late TRE and active control groups (B, E, H), and early and late TRE groups (C, F, I). Data layers include monocyte transcripts (A-C), lipid species (D-F), and metabolites (G-I). Blue and red points denote features significantly decreased or increased (FDR < 0.05). Abbreviations: Cer, ceramides; FDR, False discovery rare; SM, sphingomyelins; TAG, triacylglycerols; TRE, time-restricted eating.

**Table S1.**

| **Baseline characteristics** | **All enrolled** | **Randomized** | | |
|---|---|---|---|---|
| | | **Early TRE** | **Late TRE** | **Active control** |
| **Demographics** | **(n = 92)** | **(n = 21)** | **(n = 17)** | **(n = 19)** |
| Highest level of education | | | | |
| Secondary / high school, college | 7 (7.6%) | 1 (4.8%) | 1 (5.9%) | 1 (5.3%) |
| Apprenticeship | 15 (16.3%) | 6 (28.6%) | 0 | 4 (21.1%) |
| Professional school | 32 (34.8%) | 5 (23.8%) | 8 (47.1%) | 7 (36.8%) |
| University / applied sciences | 33 (35.9%) | 8 (38.1%) | 8 (47.1%) | 5 (26.3%) |
| Other | 5 (5.4%) | 1 (4.8%) | 0 | 2 (10.5%) |
| **Lab measurements** | **(n = 86)** | **(n = 21)** | **(n = 17)** | **(n = 19)** |
| Fasting plasma glucose, mmol/l | 5.04 ± 0.46 | 5.09 ± 0.50 | 5.14 ± 0.45 | 5.16 ± 0.36 |
| in mg/dl | 90.69 ± 8.34 | 91.54 ± 9.01 | 92.44 ± 8.10 | 92.94 ± 6.43 |
| HOMA-IR | 1.94 (1.32 to 2.75) | 2.26 (1.48 to 3.32) | 2.13 (1.41 to 2.87) | 1.58 (1.31 to 2.34) |
| HbA1c[1], % | 5.15 ± 0.31 | 5.16 ± 0.35 | 5.10 ± 0.35 | 5.17 ± 0.26 |
| Lipid profile, mmol/l | | | | |
| Total cholesterol | 4.46 (3.87 to 4.86) | 4.20 (3.85 to 4.64) | 4.19 (3.84 to 4.50) | 4.70 (3.83 to 4.98) |
| HDL cholesterol | 1.44 (1.27 to 1.63) | 1.38 (1.20 to 1.48) | 1.57 (1.35 to 1.67) | 1.45 (1.23 to 1.62) |
| LDL cholesterol | 2.55 (2.08 to 3.04) | 2.49 (2.10 to 2.82) | 2.18 (1.90 to 2.79) | 2.86 (2.20 to 3.12) |
| Triglycerides | 0.84 (0.64 to 1.07) | 0.88 (0.73 to 1.32) | 0.85 (0.63 to 0.96) | 0.78 (0.52 to 1.02) |
| **Body composition by BIA** | **(n = 90)** | **(n = 21)** | **(n = 17)** | **(n = 19)** |
| Fat mass, kg | 31.4 ± 5.7 | 30.0 ± 6.3 | 32.4 ± 5.1 | 32.0 ± 5.6 |
| Fat-free mass, kg | 52.2 ± 8.9 | 50.0 ± 8.8 | 50.2 ± 6.3 | 50.9 ± 7.3 |

| Baseline characteristics | All enrolled | Randomized | | |
|---|---|---|---|---|
| | | Early TRE | Late TRE | Active control |
| **Indirect calorimetry** | **(n = 88)** | **(n = 21)** | **(n = 17)** | **(n = 19)** |
| Resting energy expenditure, kcal/d | 1549 (1389 to 1663) | 1459 (1381 to 1593) | 1585 (1462 to 1611) | 1562 (1418 to 1644) |
| Respiratory quotient | 0.84 ± 0.06 | 0.83 ± 0.06 | 0.84 ± 0.05 | 0.86 ± 0.06 |
| **Daily intake** | **(n = 90)** | **(n = 21)** | **(n = 17)** | **(n = 19)** |
| Calorie intake, kcal/d | 1751.6 ± 342.2 | 1726.4 ± 363.9 | 1725.8 ± 231.5 | 1698.9 ± 361.7 |
| Carbohydrates, g | 182.8 ± 40.9 | 173.7 ± 35.8 | 183.5 ± 29.6 | 180.1 ± 44.5 |
| Protein, g | 73.5 ± 15.0 | 76.4 ± 17.5 | 68.7 ± 9.4 | 69.8 ± 16.5 |
| Fat, g | 80.0 ± 17.4 | 79.7 ± 21.1 | 79.5 ± 11.7 | 77.5 ± 15.9 |
| Alcohol, g | 4.2 (1.5 to 8.8) | 5.8 (3.7 to 7.9) | 3.8 (1.9 to 10.9) | 3.6 (1.7 to 6.9) |
| Fibre, g | 18.0 ± 5.0 | 18.3 ± 5.2 | 19.2 ± 5.2 | 17.1 ± 4.5 |
| **Proportion of daily calorie intake** | **(n = 90)** | **(n = 21)** | **(n = 17)** | **(n = 19)** |
| Carbohydrates,% | 41.7 ± 3.7 | 40.4 ± 3.8 | 42.5 ± 3.0 | 42.2 ± 3.5 |
| Protein, % | 16.9 ± 2.4 | 17.8 ± 2.4 | 16.0 ± 1.6 | 16.5 ± 1.9 |
| Fat, % | 41.0 ± 3.1 | 41.1 ± 3.7 | 41.4 ± 2.3 | 41.3 ± 2.6 |
| Alcohol, % | 1.9 (0.6 to 3.6) | 2.2 (1.4 to 3.5) | 1.5 (0.7 to 3.7) | 1.8 (0.7 to 3.1) |
| **Self-reported sleep habits[2]** | **(n = 89)** | **(n = 21)** | **(n = 17)** | **(n = 19)** |
| Sleep duration, h | 7.0 (7.0 to 8.0) | 7.0 (7.0 to 8.0) | 7.0 (7.0 to 7.5) | 7.0 (6.5 to 7.5) |
| **Self-reported habitual activity[2]** | **(n = 60)** | **(n = 13)** | **(n = 13)** | **(n = 10)** |
| Physical activity, METs/h | 1665 (918 to 2810) | 1718 (1116 to 2754) | 1626 (1224 to 3612) | 942 (630 to 2296) |
| **Actigraphy[3]** | **(n = 85)** | **(n = 20)** | **(n = 17)** | **(n = 19)** |
| Light activity, mins | 122.8 ± 35.8 | 129.1 ± 33.8 | 109.4 ± 35.6 | 114.1 ± 26.4 |
| Moderate activity, mins | 100.5 (76.1 to 121.2) | 96.6 (74.9 to 123.9) | 91.7 (70.2 to 109.6) | 80.9 (67.5 to 105.0) |

| Baseline characteristics | All enrolled | Randomized | | |
|---|---|---|---|---|
| | | Early TRE | Late TRE | Active control |
| Vigorous activity, mins | 1.3 (0.6 to 3.1) | 2.3 (0.5 to 3.8) | 0.8 (0.5 to 1.7) | 0.7 (0.6 to 1.8) |
| Sleep onset, hh:mm | 22:53 ± 00:59 | 23:12 ± 1:03 | 22:40 ± 00:46 | 22:40 ± 00:45 |
| Wake-up time, hh:mm | 07:03 ± 00:46 | 07:07 ± 00:43 | 06:58 ± 00:28 | 07:04 ± 00:31 |
| Sleep duration, h | 7.33 (6.69 to 7.80) | 7.12 (6.35 to 7.46) | 7.46 (7.07 to 7.83) | 7.46 (7.12 to 7.74) |

**Table S1**. Baseline characteristics of all enrolled participants (left column), then per randomized intervention group (right columns). Note that the final numbers of analyzed participants per intervention vary slightly due to study withdrawals or wearable-device technical issues (see text and STAR Methods). Data are represented as mean ± SD, median (interquartile range) or number (%) as appropriate. Time measures are expressed as clock time (hh:mm), while durations are expressed as decimal hours (h). [1] HbA1c was measured in 85 individuals of the overall included participants, due to a lab technical issue. [2] Self-reported sleep duration over the previous 4 weeks was assessed using the Pittsburgh Sleep Quality Index,[1] and habitual physical activity over the previous 4 weeks using the short form of the International Physical Activity Questionnaire.[2] [3] The other activity and sleep parameters were recorded with actigraphy and analysed with the GGIR package.[3] *Related to Table 1.* Abbreviations: BIA, bioelectrical impedance analysis; HbA1c, glycated hemoglobin; h, hours; HDL, high-density lipoprotein; HOMA-IR, homeostasis model assessment of insulin resistance; LDL, low-density lipoprotein; METs, metabolic equivalents of task; mins, minutes; TRE, time-restricted eating.

**Table S2.**

| Changes in weight and body composition | Change from baseline | | | Early TRE vs.<br>Active control | | Late TRE vs.<br>Active control | | Early TRE vs.<br>Late TRE | |
|---|---|---|---|---|---|---|---|---|---|
| | **Early TRE** | **Late TRE** | **Active control** | **Mean difference (95% CI)** | **p-value** | **Mean difference (95% CI)** | **p-value** | **Mean difference (95% CI)** | **p-value** |
| **Clinical measurements** | **(n = 19)** | **(n = 17)** | **(n = 18)** | | | | | | |
| Weight, kg | -4.51 ± 3.54 | -1.22 ± 2.33 | -0.53 ± 1.67 | -4.27 (-6.16, -2.38) | <0.001<br>0.001 | -0.66 (-2.10, 0.78) | 0.36<br>0.55 | -3.36 (-5.52, -1.21) | 0.003<br>0.02 |
| Relative weight change % | -5.55 ± 4.25 | -1.62 ± 2.80 | -0.62 ± 1.98 | -4.93 (-7.22, -2.63) | <0.001<br>0.002 | -1.00 (-2.71, 0.71) | 0.24<br>0.44 | -3.93 (-6.47, -1.39) | 0.003<br>0.02 |
| Body mass index, $kg/m^2$ | -1.63 ± 1.27 | -0.48 ± 0.82 | -0.18 ± 0.59 | -1.45 (-2.15, -0.75) | <0.001<br>0.002 | -0.31 (-0.83, 0.21) | 0.23<br>0.44 | -1.15 (-1.92, -0.39) | 0.004<br>0.03 |
| Waist circumference, cm | -3.90 ± 4.42 | -0.14 ± 4.04 | -0.75 ± 3.17 | -3.32 (-5.89, -0.74) | 0.01<br>0.049 | -0.06 (-2.55, 2.42) | 0.96<br>0.96 | -3.22 (-6.02, -0.41) | 0.03<br>0.08 |
| Hip circumference, cm | -3.80<br>(-5.15 to -2.10) | -0.20<br>(-2.30 to 2.00) | -0.60<br>(-4.58 to 3.85) | -4.07 (-7.75, -0.38) | 0.03<br>0.09 | 0.46 (-3.06, 3.97) | 0.79<br>0.85 | -4.26 (-7.55, -0.97) | 0.01<br>0.049 |
| Waist to hip ratio | -0.00 ± 0.03 | -0.00 ± 0.03 | -0.00 ± 0.04 | 0.00 (-0.02, 0.03) | 0.72<br>0.83 | -0.00 (-0.03, 0.02) | 0.81<br>0.85 | 0.01 (-0.01, 0.03) | 0.55<br>0.75 |
| **Body composition by DXA**[1] | **(n = 18)** | **(n = 17)** | **(n = 18)** | | | | | | |
| Fat mass, kg | -2.90<br>(-3.61 to -1.74) | -1.37<br>(-2.16 to 0.24) | 0.02<br>(-1.63 to 1.21) | -3.04 (-4.78, -1.29) | 0.001 | -1.05 (-2.53, 0.44) | 0.16 | -1.93 (-3.72, -0.14) | 0.04 |
| Visceral fat mass, g | -48.0<br>(-106.8 to -20.8) | -30.0<br>(-62.0 to 1.0) | -26.0<br>(-65.8 to 36.3) | -23.7 (-77.6, 30.2) | 0.38<br>0.57 | -29.4 (-87.1, 28.2) | 0.31<br>0.49 | 17.1 (-45.9, 80.1) | 0.58<br>0.75 |
| Lean mass, kg | -1.69 ± 1.89 | -0.32 ± 1.18 | -0.06 ± 1.56 | -1.69 (-2.88, -0.50) | 0.007<br>0.035 | -0.17 (-1.13, 0.79) | 0.72<br>0.83 | -1.36 (-2.46, -0.26) | 0.02<br>0.06 |
| Bone mass, kg | 0.03<br>(-0.02 to 0.04) | -0.00<br>(-0.03 to 0.02) | 0.01<br>(-0.01 to 0.03) | -0.10 (-0.21, -0.00) | 0.045<br>0.11 | -0.05 (-0.20, 0.10) | 0.51<br>0.72 | -0.08 (-0.22, 0.06) | 0.28<br>0.46 |

| Changes in weight and body composition | Change from baseline | | | Early TRE vs. Active control | | Late TRE vs. Active control | | Early TRE vs. Late TRE | |
|---|---|---|---|---|---|---|---|---|---|
| | Early TRE | Late TRE | Active control | Mean difference (95% CI) | p-value | Mean difference (95% CI) | p-value | Mean difference (95% CI) | p-value |
| **Relative change in body composition by DXA[1]** | **(n = 18)** | **(n = 17)** | **(n = 18)** | | | | | | |
| Fat mass % | -1.41<br>(-2.31 to -0.22) | -1.17<br>(-2.13 to 0.79) | 0.30<br>(-1.18 to 1.48) | -1.96 (-3.77, -0.15) | 0.035<br>0.09 | -0.82 (-2.28, 0.65) | 0.27<br>0.46 | -1.02 (-2.75, 0.72) | 0.24<br>0.44 |
| Visceral fat mass % | -0.03<br>(-0.11 to 0.01) | -0.03<br>(-0.06 to 0.00) | -0.04<br>(-0.09 to 0.06) | 0.02 (-0.05, 0.09) | 0.58<br>0.75 | -0.03 (-0.10, 0.04) | 0.41<br>0.59 | 0.06 (-0.02, 0.14) | 0.16<br>0.33 |
| Lean mass % | 1.10<br>(0.23 to 1.71) | 0.29<br>(-0.48 to 1.13) | -0.00<br>(-1.10 to 1.78) | 1.15 (-0.16, 2.45) | 0.08<br>0.20 | 0.16 (-0.93, 1.24) | 0.77<br>0.85 | 1.01 (-0.16, 2.18) | 0.09<br>0.20 |
| Bone mass % | 0.18<br>(0.08 to 0.29) | 0.07<br>(-0.02 to 0.11) | 0.05<br>(-0.00 to 0.09) | 0.03 (-0.11, 0.18) | 0.65<br>0.81 | -0.01 (-0.19, 0.16) | 0.87<br>0.88 | 0.03 (-0.16, 0.23) | 0.72<br>0.83 |
| **Body composition by BIA** | **(n = 19)** | **(n = 17)** | **(n = 18)** | | | | | | |
| Fat mass, kg | -2.94 ± 2.66 | -0.90 ± 1.83 | -0.75 ± 1.60 | -2.12 (-3.69, -0.55) | 0.01<br>0.04 | -0.15 (-1.39, 1.08) | 0.80<br>0.85 | -1.93 (-3.60, -0.26) | 0.025<br>0.08 |
| Fat-free mass, kg | -1.57 ± 1.25 | -0.32 ± 0.96 | 0.22 ± 0.88 | -1.81 (-2.56, -1.05) | <0.001<br>0.001 | -0.49 (-1.12, 0.14) | 0.12<br>0.26 | -1.25 (-2.04, -0.45) | 0.003<br>0.02 |

**Table S2**. Body composition and clinical measurements within each randomized group and comparisons of these changes between groups among participants who completed the study. Data are represented as mean ± SD, median (interquartile range) or mean (95%CI) as appropriate. Two p-values are reported: (top) unadjusted p-value, (bottom) p-value adjusted for multiple testing with the Benjamini-Hochberg procedure, except for the primary outcome. [1] Body composition was assessed by DXA in 18 out of 19 participants of the early TRE group, due to limited availability of DXA measurement within the required timeframe. *Related to Figure 2*. Abbreviations: BIA, bioelectrical impedance analysis; CI, confidence interval; DXA, dual energy X-ray absorptiometry; TRE, time-restricted eating.

**Table S3.**

| Changes in metabolic health, activity and sleep parameters | Change from baseline | | | Early TRE vs. Active control | | Late TRE vs. Active control | | Early TRE vs. Late TRE | |
|---|---|---|---|---|---|---|---|---|---|
| | Early TRE | Late TRE | Active control | Mean difference (95% CI) | p-value | Mean difference (95% CI) | p-value | Mean difference (95% CI) | p-value |
| **Clinical measurements** | **(n = 19)** | **(n = 17)** | **(n = 18)** | | | | | | |
| Systolic BP, mmHg | -3.9 ± 8.0 | -0.3 ± 5.2 | -3.1 ± 11.4 | -1.4 (-6.1, 3.2) | 0.54<br>0.84 | 0.2 (-4.5, 4.9) | 0.94<br>0.99 | -2.3 (-6.3, 1.8) | 0.27<br>0.77 |
| Diastolic BP, mmHg | -3.0 ± 6.3 | 0.9 ± 4.6 | -1.1 ± 8.4 | -1.8 (-5.6, 2.0) | 0.33<br>0.83 | 0.9 (-3.2, 5.0) | 0.65<br>0.84 | -2.9 (-6.3, 0.5) | 0.09<br>0.50 |
| Fasting plasma glucose, mmol/l | -0.33 ± 0.43 | -0.22 ± 0.26 | -0.06 ± 0.30 | -0.27 (-0.52, -0.03) | 0.03<br>0.30 | -0.17 (-0.37, 0.03) | 0.10<br>0.51 | -0.09 (-0.32, 0.14) | 0.44<br>0.83 |
| in mg/dl | -5.87 ± 7.70 | -4.02 ± 4.72 | -1.00 ± 5.44 | -4.90 (-9.29, -0.50) | | -3.06 (-6.74, 0.61) | | -1.62 (-5.81, 2.57) | |
| HOMA-IR | -1.19 ± 1.13 | -0.55 ± 0.49 | -0.14 ± 0.67 | -0.57 (-1.09, -0.05) | 0.03<br>0.30 | -0.34 (-0.73, 0.05) | 0.08<br>0.50 | -0.27 (-0.72, 0.17) | 0.22<br>0.68 |
| HbA1c[1], % | -0.05 ± 0.27 | 0.03 ± 0.23 | -0.01 ± 0.22 | -0.04 (-0.19, 0.11) | 0.57<br>0.84 | 0.04 (-0.13, 0.20) | 0.65<br>0.84 | -0.07 (-0.23, 0.10) | 0.42<br>0.83 |
| Lipid profile, mmol/l | | | | | | | | | |
| Total cholesterol | -0.08 ± 0.46 | -0.06 ± 0.40 | -0.25 ± 0.31 | 0.16 (-0.12, 0.43) | 0.25<br>0.75 | 0.18 (-0.08, 0.43) | 0.17<br>0.61 | -0.02 (-0.32, 0.28) | 0.89<br>0.97 |
| HDL cholesterol | -0.00 ± 0.11 | -0.05 ± 0.16 | -0.01 ± 0.13 | 0.00 (-0.08, 0.09) | 0.98<br>0.99 | -0.02 (-0.12, 0.09) | 0.72<br>0.89 | 0.01 (-0.09, 0.11) | 0.84<br>0.95 |
| LDL cholesterol | 0.03 ± 0.40 | 0.03 ± 0.31 | -0.18 ± 0.32 | 0.18 (-0.08, 0.43) | 0.16<br>0.61 | 0.16 (-0.06, 0.39) | 0.15<br>0.61 | 0.00 (-0.26, 0.26) | 0.99<br>0.99 |
| Triglycerides[2] | -0.13<br>(-0.32 to -0.03) | -0.09<br>(-0.24 to 0.02) | -0.10<br>(-0.28 to 0.03) | 4.5 (-12.8, 25.1) | 0.62<br>0.84 | 2.2 (-15.3, 23.2) | 0.82<br>0.95 | 0.4 (-15.7, 19.5) | 0.97<br>0.99 |

| Changes in metabolic health, activity and sleep parameters | Change from baseline | | | Early TRE vs. Active control | | Late TRE vs. Active control | | Early TRE vs. Late TRE | |
|---|---|---|---|---|---|---|---|---|---|
| | Early TRE | Late TRE | Active control | Mean difference (95% CI) | p-value | Mean difference (95% CI) | p-value | Mean difference (95% CI) | p-value |
| **Indirect calorimetry** | **(n = 19)** | **(n = 17)** | **(n = 18)** | | | | | | |
| Resting energy expenditure, kcal/d | -56 (-139 to -11) | -82 (-124 to -19) | -32 (-112 to 49) | -55.7 (-144.1, 32.6) | 0.21 0.68 | -30.6 (-123.7, 62.5) | 0.51 0.84 | -21.3 (-110.7, 68.2) | 0.63 0.84 |
| Respiratory quotient | -0.04 ± 0.06 | -0.00 ± 0.08 | -0.04 ± 0.08 | -0.03 (-0.06, 0.01) | 0.14 0.61 | 0.01 (-0.03, 0.05) | 0.52 0.84 | -0.04 (-0.08, -0.01) | 0.007 0.14 |
| **Self-reported sleep habits[3]** | **(n = 18)** | **(n = 17)** | **(n = 18)** | | | | | | |
| Sleep duration, h | 0.00 (-0.88 to 0.00) | 0.00 (-0.50 to 1.00) | 0.00 (-0.88 to 0.50) | 0.71 (-0.89, 2.30) | 0.37 0.83 | 0.20 (-0.32, 0.72) | 0.44 0.83 | 0.42 (-1.20, 2.04) | 0.60 0.84 |
| **Self-reported habitual activity[3]** | **(n = 9)** | **(n = 13)** | **(n = 7)** | | | | | | |
| Physical activity, METs/h | 951.8 ± 2284.6 | -570.6 ± 1963.8 | -122.6 ± 1358.4 | 49.3 (-67.7, 589.3) | 0.58 0.84 | 25.1 (-63.2, 324.9) | 0.70 0.89 | 52.0 (-51.4, 375.0) | 0.45 0.83 |
| **Actigraphy[4]** | **(n = 18)** | **(n = 17)** | **(n = 17)** | | | | | | |
| Light activity, mins | -2.38 ± 23.75 | 1.47 ± 27.07 | 5.81 ± 17.80 | -3.6 (-14.7, 8.9) | 0.54 0.84 | -5.1 (-17.9, 9.8) | 0.47 0.84 | 1.5 (-13.5, 19.0) | 0.85 0.95 |
| Moderate activity, mins | -5.04 (-13.20 to 4.49) | -6.15 (-21.52 to 2.60) | 11.02 (4.85 to 22.48) | -18.8 (-27.7, -8.9) | <0.001 0.05 | -22.6 (-34.8, -8.3) | 0.004 0.12 | 7.1 (-8.3, 25.1) | 0.37 0.83 |
| Vigorous activity, mins | 0.06 (-0.27 to 0.45) | 0.09 (-0.62 to 0.63) | 0.25 (-0.06 to 0.65) | -11.4 (-48.2, 51.6) | 0.65 0.84 | -23.4 (-61.0, 50.5) | 0.43 0.83 | 27.4 (-29.9, 131.5) | 0.41 0.83 |
| Sleep onset, hh:mm | -00:05 (-00:17 to 00:33) | -00:01 (-00:08 to 00:31) | 00:05 (-00:14 to 00:37) | -00:04 (-00:43, 00:35) | 0.84 0.95 | -00:02 (-00:28, 00:25) | 0.90 0.97 | -00:03 (-00:39, 00:32) | 0.85 0.95 |
| Wake-up time, hh:mm | -00:16 (-00:43 to 00:23) | 00:19 (-00:10 to 00:31) | -00:04 (-00:31 to 00:25) | -00:11 (-00:40, 00:18) | 0.44 0.83 | 00:17 (-00:06, 00:40) | 0.14 0.61 | -00:29 (-01:01, 00:03) | 0.07 0.50 |

| Changes in metabolic health, activity and sleep parameters | Change from baseline | | | Early TRE vs. Active control | | Late TRE vs. Active control | | Early TRE vs. Late TRE | |
|---|---|---|---|---|---|---|---|---|---|
| | **Early TRE** | **Late TRE** | **Active control** | **Mean difference (95% CI)** | **p-value** | **Mean difference (95% CI)** | **p-value** | **Mean difference (95% CI)** | **p-value** |
| Sleep duration, h | -0.23 ± 0.52 | 0.21 ± 0.45 | -0.09 ± 0.39 | -0.15 (-0.49, 0.20) | 0.39<br>0.83 | 0.30 (-0.00, 0.61) | 0.051<br>0.42 | -0.42 (-0.78, -0.06) | 0.02<br>0.30 |

**Table S3**. Changes in metabolic health, indirect calorimetry, activity and sleep parameters within each randomized group and comparisons of these changes between groups. Data are represented as mean ± SD, median (interquartile range) or mean (95%CI) as appropriate. Time measures are expressed as clock time (hh:mm), while durations are expressed as decimal hours (h). Two p-values are reported: (top) unadjusted p-value, (bottom) p-value adjusted for multiple testing with the Benjamini-Hochberg procedure. [1] HbA1c was measured in 17 participants of the active control group, due to a lab technical issue. [2] These variables were log-transformed for analysis. Their results are presented as percent differences. [3] Self-reported sleep duration over the previous 4 weeks was assessed using the Pittsburgh Sleep Quality Index,[1] and habitual physical activity over the previous 4 weeks using the short form of the International Physical Activity Questionnaire[2]. [4] The other activity and sleep parameters in this table were recorded with actigraphy and analysed with the GGIR package.[3] *Related to Figure 2*. Abbreviations: BP, blood pressure; CI, confidence interval; d, day; HbA1c, glycated hemoglobin; h, hours; HDL, high-density lipoprotein; LDL, low-density lipoprotein; METs, metabolic equivalents of task; mins, minutes; TRE, time-restricted eating.

**Table S4.**

| Changes in calorie and macronutrient intake | Change from baseline | | | Early TRE vs. Active control | | Late TRE vs. Active control | | Early TRE vs. Late TRE | |
|---|---|---|---|---|---|---|---|---|---|
| | Early TRE (n = 19) | Late TRE (n = 17) | Active control (n = 18) | Mean difference (95% CI) | p-value | Mean difference (95% CI) | p-value | Mean difference (95% CI) | p-value |
| **Daily intake** | | | | | | | | | |
| Calorie intake, kcal/d | -399.3 ± 285.8 | -196.7 ± 239.1 | -132.7 ± 368.6 | -263.1 (-472.8, -53.4) | 0.02<br>0.10 | -56.9 (-262.1, 148.2) | 0.58<br>0.74 | -205.5 (-370.4, -40.5) | 0.02<br>0.10 |
| Carbohydrates, g | -31.8<br>(-49.8 to -25.7) | -29.2<br>(-39.4 to -15.0) | -15.1<br>(-28.0 to -4.9) | -22.4 (-44.7, 0.0) | 0.050<br>0.15 | -3.6 (-25.6, 18.5) | 0.74<br>0.82 | -18.8 (-35.9, -1.7) | 0.03<br>0.14 |
| Protein, g | -17.8 ± 15.5 | -6.7 ± 9.2 | 0.4 ± 16.3 | -15.9 (-26.0, -5.8) | 0.003<br>0.03 | -7.8 (-16.5, 0.9) | 0.08<br>0.18 | -7.6 (-15.6, 0.5) | 0.06<br>0.16 |
| Fat, g | -18.9 ± 16.1 | -9.1 ± 12.8 | -6.6 ± 21.0 | -11.9 (-23.6, -0.2) | 0.047<br>0.15 | -1.7 (-13.2, 9.7) | 0.76<br>0.82 | -10.0 (-18.9, -1.0) | 0.03<br>0.14 |
| Fibre, g | -4.6<br>(-6.9 to -3.4) | -2.9<br>(-4.8 to -0.04) | -0.7<br>(-2.9 to 0.2) | -5.3 (-8.1, -2.5) | <0.001<br>0.01 | -2.1 (-5.3, 1.2) | 0.20<br>0.40 | -3.3 (-5.1, -1.4) | <0.001<br>0.01 |
| Alcohol, g | -0.5<br>(-2.8 to 1.3) | -0.0<br>(-1.2 to 1.6) | 0.6<br>(-1.3 to 3.5) | -2.6 (-5.4, 0.1) | 0.057<br>0.15 | -2.1 (-5.7, 1.5) | 0.24<br>0.42 | -0.6 (-3.2, 1.9) | 0.62<br>0.74 |
| **Proportion of daily calorie intake** | | | | | | | | | |
| Carbohydrates,% | 0.8 ± 4.5 | -0.6 ± 2.1 | -1.2 ± 6.5 | 1.0 (-2.6, 4.5) | 0.59<br>0.74 | 0.8 (-2.5, 4.1) | 0.62<br>0.74 | 0.7 (-1.8, 3.2) | 0.56<br>0.74 |
| Protein, % | -0.7<br>(-1.4 to 1.4) | 0.4<br>(-0.2 to 1.0) | 1.5<br>(-0.0 to 2.8) | -1.3 (-3.0, 0.5) | 0.16<br>0.33 | -1.6 (-3.3, -0.01) | 0.048<br>0.15 | 0.3 (-0.9, 1.5) | 0.58<br>0.74 |
| Fat, % | -0.3<br>(-3.6 to 3.2) | 0.5<br>(-0.6 to 1.5) | -0.9<br>(-3.4 to 0.7) | 0.2 (-2.2, 2.5) | 0.89<br>0.92 | 0.6 (-1.7, 2.9) | 0.61<br>0.74 | -0.4 (-2.5, 1.8) | 0.74<br>0.82 |
| Alcohol, % | 0.4<br>(-0.3 to 0.9) | 0.1<br>(-0.4 to 1.4) | 0.5<br>(-0.1 to 1.7) | -0.8 (-2.1, 0.5) | 0.21<br>0.40 | -0.8 (-2.3, 0.7) | 0.27<br>0.46 | 0.04 (-1.1, 1.2) | 0.94<br>0.94 |

**Table S4**. Calorie intake and macronutrient consumption as recorded with the smartphone food diary app MyFoodRepo in each randomization group, then compared between groups. Data are represented as mean ± SD, median (interquartile range) or mean (95%CI) as appropriate. Two p-values are reported: (top) unadjusted p-value, (bottom) p-value adjusted for multiple testing with the Benjamini-Hochberg procedure. *Related to Figure 3, Figure S3*. Abbreviations: CI, confidence interval; d, day; TRE, time-restricted eating.

**Table S5.**

| Changes in glucose variability | Change from baseline | | | Early TRE vs. Active control | | Late TRE vs. Active control | | Early TRE vs. Late TRE | |
|---|---|---|---|---|---|---|---|---|---|
| | Early TRE | Late TRE | Active control | Mean difference (95% CI) | p-value | Mean difference (95% CI) | p-value | Mean difference (95% CI) | p-value |
| **CGM metrics** | **(n = 16)** | **(n = 13)** | **(n = 17)** | | | | | | |
| Mean glucose, mmol/l | -0.03 ± 0.44 | -0.28 ± 0.53 | 0.03 ± 0.64 | 0.02 (-0.30, 0.33) | 0.90<br>0.90 | -0.24 (-0.62, 0.14) | 0.20<br>0.27 | 0.25 (-0.11, 0.61) | 0.16<br>0.24 |
| Coefficient of variation (CV), % | 2.35<br>(1.36 to 4.34) | -1.48<br>(-2.84 to -0.71) | -0.28<br>(-0.97 to 1.81) | 2.62 (1.14, 4.09) | 0.001<br>0.004 | -0.12 (-1.86, 1.63) | 0.89<br>0.90 | 2.92 (0.72, 5.12) | 0.01<br>0.02 |
| MAGE, mmol/l | 0.28 ± 0.18 | -0.16 ± 0.35 | 0.01 ± 0.29 | 0.26 (0.10, 0.43) | 0.003<br>0.009 | -0.12 (-0.34, 0.10) | 0.28<br>0.34 | 0.40 (0.19, 0.60) | <0.001<br>0.003 |
| Standard deviation, mmol/l | 0.12 ± 0.11 | -0.07 ± 0.11 | 0.01 ± 0.12 | 0.12 (0.04, 0.19) | 0.004<br>0.009 | -0.06 (-0.13, 0.02) | 0.13<br>0.23 | 0.18 (0.10, 0.27) | <0.001<br>0.002 |
| **Glucose dynamics metrics** | **(n = 14)** | **(n = 12)** | **(n = 15)** | | | | | | |
| Glucose 24h rhythm | | | | | | | | | |
| Baseline, mmol/l | -0.12 ± 0.45 | -0.33 ± 0.61 | 0.05 ± 0.62 | -0.06 (-0.41, 0.28) | 0.71<br>0.85 | -0.26 (-0.65, 0.14) | 0.20<br>0.30 | 0.21 (-0.19, 0.61) | 0.28<br>0.38 |
| Amplitude[1], mmol/l | 0.28<br>(0.06 to 0.42) | 0.03<br>(-0.10 to 0.25) | -0.03<br>(-0.14 to 0.16) | 67.0 (23.4, 126.1) | 0.002<br>0.02 | 2.4 (-37.5, 67.9) | 0.92<br>0.98 | 62.7 (-0.5, 166.1) | 0.052<br>0.14 |
| Postprandial glucose excursions | | | | | | | | | |
| Meal height, mmol/l | 0.11 ± 0.19 | -0.12 ± 0.15 | 0.01 ± 0.09 | 0.12 (-0.00, 0.23) | 0.054<br>0.14 | -0.08 (-0.17, 0.01) | 0.06<br>0.14 | 0.20 (0.06, 0.33) | 0.008<br>0.046 |
| Half life[1], hours | 0.15<br>(0.09 to 0.29) | 0.09<br>(0.03 to 0.24) | -0.08<br>(-0.18 to 0.20) | 10.6 (-1.4, 24.2) | 0.08<br>0.14 | 14.2 (-1.0, 31.8) | 0.07<br>0.14 | -0.1 (-13.0, 14.6) | 0.98<br>0.98 |

**Table S5**. CGM and glucose dynamics metrics in each randomization group, then compared between groups. Data are represented as mean ± SD, median (interquartile range) or mean (95%CI) as appropriate. Sample sizes differ from other analyses because only participants with at least 48 hours of CGM data were included. Two p-values are reported: (top) unadjusted p-value, (bottom) p-value adjusted for multiple testing with the Benjamini-Hochberg procedure, which were calculated separately for CGM metrics and glucose dynamics metrics. [1] These variables were log-transformed for analysis. Their results are presented as percent differences. *Related to Figure 4-5*. Abbreviations: CGM, continuous glucose monitoring; CI, confidence interval; CV, coefficient of variation; MAGE, mean amplitude of glycemic excursions; TRE, time-restricted eating.
